# Co-expression-wide association studies link genetically regulated interactions with complex traits

**DOI:** 10.1101/2024.10.02.24314813

**Authors:** Mykhaylo M. Malakhov, Wei Pan

**Affiliations:** Division of Biostatistics and Health Data Science, School of Public Health, University of Minnesota, Minneapolis, MN, USA

## Abstract

Transcriptome- and proteome-wide association studies (TWAS/PWAS) have proven successful in prioritizing genes and proteins whose genetically regulated expression modulates disease risk, but they ignore potential co-expression and interaction effects. To address this limitation, we introduce the co-expression-wide association study (COWAS) method, which can identify pairs of genes or proteins whose genetically regulated co-expression is associated with complex traits. COWAS first trains models to predict expression and co-expression from genetic variation, and then tests for association between imputed co-expression and the trait of interest while also accounting for direct effects from each exposure. We applied our method to plasma proteomic concentrations from the UK Biobank, identifying dozens of interacting protein pairs associated with cholesterol levels, Alzheimer’s disease, and Parkinson’s disease. Notably, our results demonstrate that co-expression between proteins may affect complex traits even if neither protein is detected to influence the trait when considered on its own. We also show how COWAS can help to disentangle direct and interaction effects, providing a richer picture of the molecular networks that mediate genetic effects on disease outcomes.

## Introduction

Translating genetic associations into knowledge of causal genes and proteins is a central problem in genetic epidemiology. Although genome-wide association studies (GWAS) can rapidly identify the single nucleotide polymorphisms (SNPs) and genetic loci associated with any measurable phenotype, most of the significant GWAS hits for complex traits fall outside of protein-coding regions and are thought to affect the phenome through regulatory pathways [1–6]. A popular approach for aggregating these regulatory effects into interpretable gene-level functional units is the transcriptome-wide association study (TWAS) method [7, 8]. TWAS is a two-stage framework that first trains a model to predict gene expression levels from genetic variation, thereby estimating the genetically regulated component of expression, and then tests for association between imputed expression and the trait of interest. Although most commonly applied to gene expression data, TWAS can be used with any heritable molecular phenotype. For example, proteome-wide association studies (PWAS) identify disease-relevant proteins by applying the two-stage TWAS framework to proteomic concentrations [9–11].

Many innovative methodological extensions to TWAS and PWAS have been developed since their initial introductions [12–20], with applications spanning hundreds of outcome traits [21–27]. All existing TWAS/PWAS methods, however, have a major limitation: they fail to account for correlations or interactions among the functional units being studied. In standard TWAS approaches, each gene or protein is considered independently of the rest. This marginal assumption is mathematically simple and provides for a straightforward implementation of the method, but it is biologically implausible. Moreover, discounting interaction effects in TWAS may lead to a loss of statistical power and missed biological insights when considering molecular drivers that primarily affect complex traits through synergistic pathways.

Recent methods have partially addressed the marginal limitation in TWAS by fine-mapping candidate TWAS genes to separate the effects of multiple correlated exposures [28–30]. These methods can tease out the likely causal genes within a larger set of co-expressed genes by conditioning each gene on the others. However, they do not model the genetic regulation of co-expression and cannot be used to infer the impact of gene–gene or protein–protein interactions on the outcome trait. In a separate line of research, protein–protein interaction (PPI) networks have been used to aid in the interpretation of PWAS findings [31]. Such use of PPI networks, however, still relies on the results of testing each protein individually for association with disease, and only utilizes evidence of interactions to cluster those marginal associations. Thus, no existing approaches are able to elucidate the extent to which co-expression and interactions among molecular phenotypes mediate genetic effects on complex traits.

The importance of epistasis, co-expression, and PPIs in complex disease pathogenesis has been well established and is the subject of extensive research despite the challenges of ascertaining interaction effects from genomic data [32–35]. An increasing burden of evidence also highlights the role of genetic variation in regulating gene–gene and protein–protein interactions. For example, single-cell RNA sequencing data has enabled the detection of genetic variants that significantly alter co-expression relationships [36]. More recently, a pan-cancer study demonstrated that point mutations correlate with altered, tumor-specific PPIs and can rewire interaction networks [37]. Other work used gene co-expression networks to link cancer driver genes to cancer GWAS genes, showing that common genetic variants are involved in the regulation of co-expression networks [38]. More generally, large-scale sequencing studies have established that both germline and somatic mutations are responsible for widespread perturbations of PPI networks in human diseases [39]. Such evidence suggests that it should be possible to predict the effects of genetic variation on gene or protein co-expression, and to consequently assess the association between genetically regulated co-expression and disease.

In this paper we introduce the co-expression-wide association study (COWAS) method to identify interacting genes or proteins whose genetic component of co-expression is associated with complex traits. COWAS analyzes pairs of co-expressed molecular exposures, first imputing their genetically regulated expression and co-expression, and then jointly testing for both direct effects and interaction effects on the outcome trait. We also extend the association testing stage of COWAS to a summary statistics setting, making it easy to apply our method to any trait of interest for which summary-level GWAS data are available.

We applied COWAS to plasma proteomic concentrations from the UK Biobank (UKB) [40, 41] and GWAS datasets for three complex traits [42–45]. We first trained imputation models for pairs of proteins with known PPIs, and then tested each well-imputed pair for association with low-density lipoprotein (LDL) cholesterol, Alzheimer’s disease (AD), and Parkinson’s disease (PD). The latter two traits were chosen because of our scientific interest in the genetic basis of neurodegenerative disorders, while LDL cholesterol was considered as an example of a quantitative trait for which COWAS would be especially well-powered due to the availability of very large GWAS cohorts. Our results demonstrate that COWAS can successfully identify protein pairs whose co-expression impacts complex traits, while at the same time disentangling their direct and interaction effects. Our approach also increases power relative to standard PWAS analyses, leading to the discovery of disease-relevant proteins that were missed by PWAS. Notably, we show that co-expression between proteins may affect disease risk even if neither protein significantly influences the disease when considered on its own. Overall, our contribution provides a novel framework for studying the effects of genetically regulated co-expression on complex traits, facilitating interrogation of the phenotypic consequences of gene–gene and protein–protein interactions using GWAS summary statistics.

## Results

### Overview of COWAS

The co-expression-wide association study (COWAS) method prioritizes pairs of interacting genes or proteins whose genetically regulated expression or co-expression is significantly associated with a complex trait. Note that COWAS can be applied to either gene expression or protein expression data, but since our application concerns the proteome, we will primarily refer to protein expression throughout the rest of the paper.

The key motivation behind our approach is the observation that genetic variation modulates not only protein expression, but also protein co-expression (Figure 1a) [37–39, 46–48]. We refer to genetic variants associated with co-expression as co-expression quantitative trait loci (coQTLs) [36], analogously to how variants associated with gene expression are termed expression quantitative trait loci (eQTLs) and variants associated with protein expression are termed protein quantitative trait loci (pQTLs). A variant can belong to more than one of these xQTL classes, and we assume that a coQTL is most likely also an eQTL or a pQTL. Furthermore, we consider co-expression to be a proxy for interaction effects. Although gene–gene and protein–protein interactions are not directly measured in large biobank studies such as the UKB, co-variation of protein abundance is an accurate proxy for PPIs because interacting protein pairs are known to be highly co-expressed [37, 49]. COWAS leverages pQTL data to learn the patterns of genetic regulation underlying protein expression and co-expression, and ultimately estimates the direct and interaction effects of genetically regulated expression on a complex trait of interest (Figure 1b).

**Fig. 1.**
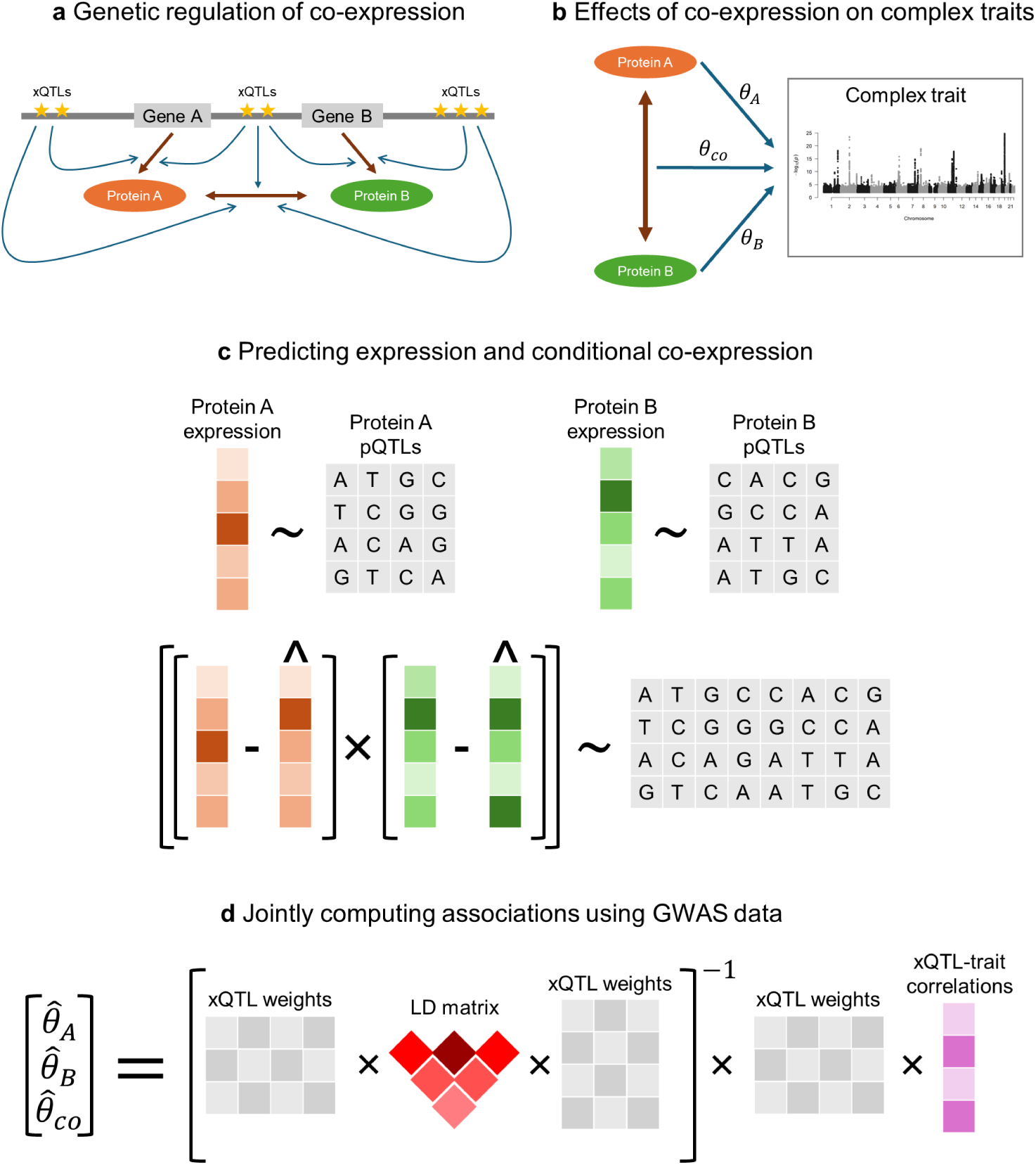
Overview of the COWAS framework. **a**, Genes A and B code for proteins A and B, which interact with each other. The transcription, translation, and interaction processes are regulated by eQTLs, pQTLs, and coQTLs, respectively, which may overlap and are collectively denoted as xQTLs. **b**, Proteins A and B may have direct effects on a complex trait (*θ_A_* and *θ_B_*, respectively), but they may also impact the trait through their interactions with each other (*θco*). **c**, The training stage of COWAS involves first building models to impute the genetically regulated expression levels of each protein from pQTL genotypes, and then building a third model to impute their genetically regulated co-expression. **d**, The testing stage of COWAS involves jointly estimating direct and interaction effects on a complex trait of interest using fitted model weights from the training stage, a linkage disequilibrium reference panel, and GWAS summary statistics for the outcome trait.

The COWAS framework is comprised of a training stage (Figure 1c) and a testing stage (Figure 1d). The training stage must be performed using individual-level genotype and expression data. First, models are trained to predict the expression levels of each protein from its pQTLs. Next, imputed expression is subtracted from measured expression to obtain expression residuals for each protein, which capture the component of expression not explained by genetic effects on its mean. Finally, a third model is trained to impute the product of these residuals from the union of all considered pQTLs. By doing so, our approach removes the component of co-expression that is explained by linear genetic effects on mean expression levels or by factors unrelated to genetics, allowing us to focus on how genetic variation modulates the amount of correlation between the two proteins. Explicitly modeling the genetic regulation of co-expression is the primary innovation of COWAS, because it enables us to incorporate the genetic component of gene or protein co-expression into an association testing framework. Note that the main version of our proposed method is not equivalent to fitting an interaction model. We alternatively considered a product-based version of COWAS that imputes the product of observed expression levels, which is more similar to a traditional interaction model. The residual-based and product-based models are both available in our R implementation of COWAS, and users can choose between them depending on the interpretation they desire. Unless otherwise stated, we used the residual-based version of COWAS to obtain the results in this paper.

The testing stage of COWAS is performed using fitted model weights from the training stage, a linkage disequilibrium (LD) reference panel, and summary-level GWAS data for the outcome trait of interest (Figure 1d). Here three effect sizes are jointly estimated: the direct effect of the first protein’s genetically regulated expression on the trait (*θ_A_*), the direct effect of the second protein’s genetically regulated expression on the trait (*θ_B_*), and the effect of their genetically regulated co-expression on the trait (*θ_co_*). Note that *θ_A_* and *θ_B_* are distinct from the marginal effects obtained through standard TWAS or PWAS, since here the three effect sizes are estimated together in a joint model. As a result, each effect size is conditional on the other two.

Several hypothesis tests can be performed with these estimated effect sizes and their standard errors. The COWAS global test determines if the protein pair has an overall effect on the outcome trait by comparing the COWAS model to a null model. This test is appropriate when the primary analysis goal is to simply identify disease-relevant protein pairs, rather than to ascertain whether or not those proteins work together to affect disease risk. As we show in subsequent sections, the COWAS global test can boost power relative to marginal TWAS/PWAS analyses of each exposure. One can also test the effect size estimates individually in order to disentangle the impact of genetically regulated co-expression from the impact of each protein’s genetically regulated expression. In particular, the COWAS interaction test determines if the parameter *θ_co_*is nonzero. This test is appropriate for checking whether co-expression has an effect on the outcome trait while accounting for direct effects from both exposures. Finally, testing the other two coefficients (*θ_A_*, *θ_B_*) reveals whether each protein has a significant direct effect on the trait while controlling for the other protein and for their co-expression. The flexibility and increased statistical power offered by COWAS enable it to identify novel disease-relevant genes or proteins and aid in the interpretation of GWAS findings. Significant COWAS protein pairs can then be visualized as a network in order to highlight groups of proteins that mediate genetic risk on the outcome trait. Although COWAS will always report the results of all available hypothesis tests, care must be taken to decide on the research question beforehand and to properly adjust the results for multiple testing.

### Accurately imputing genetically regulated co-expression

We trained COWAS models to predict protein expression and co-expression using genotype data and proteomic concentrations from unrelated White British participants in the UKB Pharma Proteomics Project [41]. After quality control, we retained 2,833 proteins coded by autosomal genes. Since training imputation models for each of the 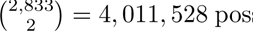 possible protein pairs would have been computationally infeasible, we excluded pairs that are unlikely to be co-expressed by restricting our analysis to pairs with experimental evidence of PPIs [50]. Model training will generally fail for protein pairs that are not co-expressed to begin with, so excluding such pairs greatly reduces the number of models that need to be trained and tested without significantly sacrificing important signals. Moreover, excluding those pairs does not introduce selection bias because experimental evidence of PPIs is independent of any downstream traits. In total, we trained COWAS models using UKB genotypes and protein abundance levels for 26,433 protein pairs, after normalizing and adjusting for standard covariates.

To ensure that COWAS can accurately predict genetically regulated co-expression, we explored the out-of-sample imputation performance of several regression methods (Figure 2). We considered penalized linear regression models with either an elastic net penalty, a lasso penalty, or a ridge penalty. For each of these three model types, we pre-screened genetic variants to reduce model training time by selecting the top 100 pQTLs for each protein based either on their *P* values or on their effect sizes. Additionally, we also considered the extent to which including both local pQTLs (*cis*-pQTLs) and distant pQTLs (*trans*-pQTLs) improved model imputation performance relative to only including *cis*-pQTLs.

**Fig. 2.**
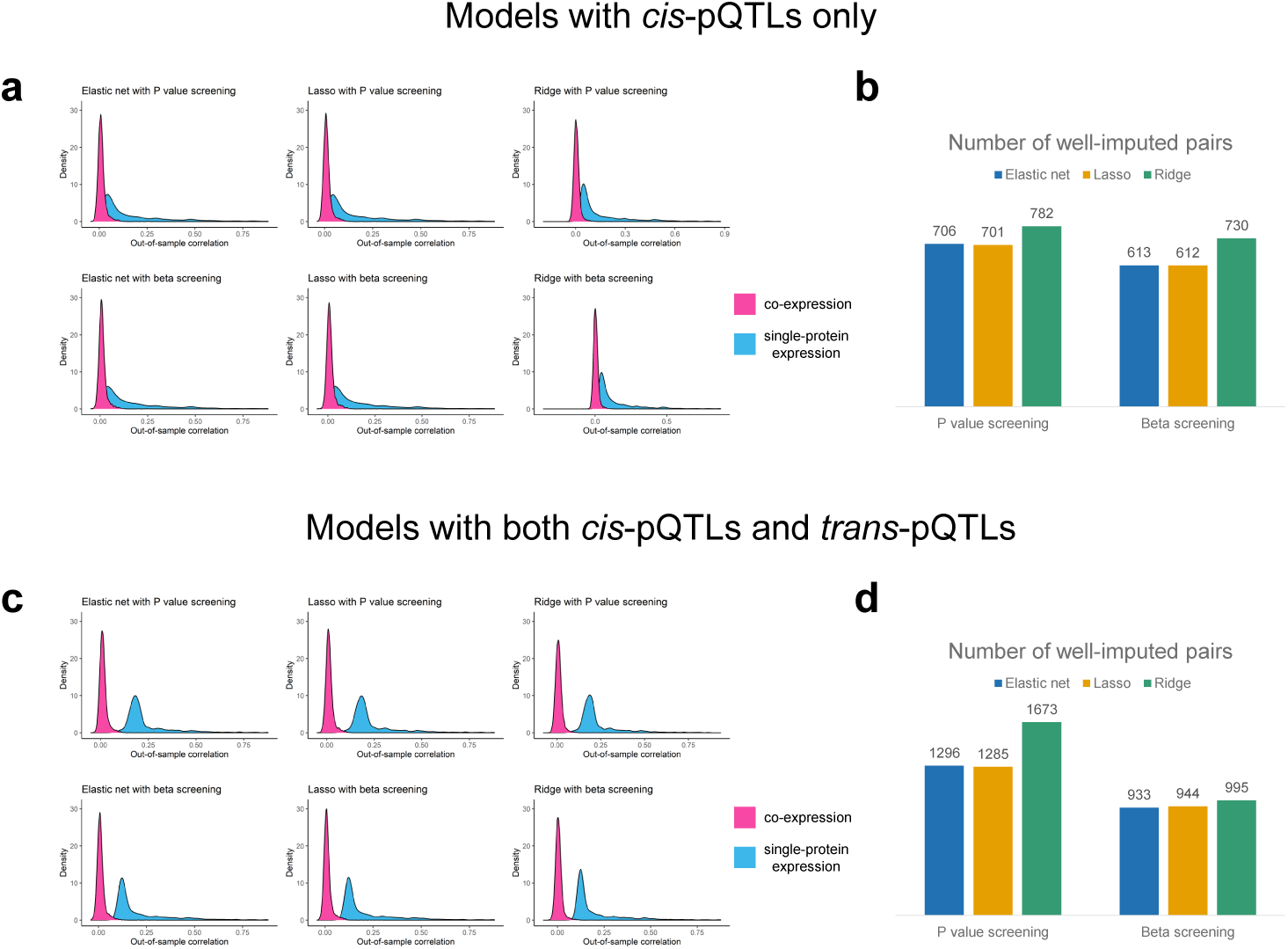
Performance metrics for COWAS models trained on UKB data. **a**,**c**, Density plots of the out-of-sample correlation between estimated and imputed co-expression (in pink), and between measured and imputed single-protein expression (in blue). **b**,**d**, Counts of the numbers of protein pairs in which all three prediction models had an out-of-sample correlation greater than 0.03. Models in **a** and **b** were trained with only *cis*-pQTLs as predictors. Models in **c** and **d** were trained with both *cis*-pQTLs and *trans*-pQTLs as predictors.

Our results show that accurate imputation is more challenging for protein co-expression than for the expression of individual proteins. Across all of the model types we considered, the median out-of-sample correlation between estimated and imputed co-expression was always lower than the median out-of-sample correlation between measured and imputed single-protein expression (Figure 2a, Figure 2c, and Supplementary Data 1). This was expected, since interaction effects are known to be more difficult to detect than main effects, with considerably larger sample sizes being needed for the same level of power or prediction quality. Interestingly, including *trans*-pQTLs in addition to *cis*-pQTLs significantly increased the imputation quality for single-protein models, but it did not have a pronounced effect on the performance of co-expression models (Figure 2a and Figure 2c). One possible explanation for this observation is that *trans*-pQTLs only weakly regulate PPIs, with the bulk of heritability in co-expression stemming from local genetic variation. However, it is also possible that *trans*-coQTLs may not overlap with *trans*-pQTLs. Since we pre-screened genetic variants based on the strength of their association with the individual proteins in each pair, the inclusion of distant variants primarily increases the number of strong pQTLs present in each model and may not necessarily increase the number of strong coQTLs. Next, we filtered the protein pairs to those in which all three imputation models yielded an out-of-sample correlation greater than 0.03 (Figure 2b and Figure 2d). Among these well-imputed pairs, lasso regression with *cis*-pQTLs pre-screened by their effect sizes achieved the highest mean out-of-sample *R*^2^ for predicting co-expression (mean *R*^2^ = 0.0038, Supplementary Data 1). On the other hand, ridge regression with both *cis*-pQTLs and *trans*-pQTLs pre-screened by their *P* values yielded the greatest number of well-imputed protein pairs (Figure 2d and Supplementary Data 1). Depending on the model type selected, between 8% and 19% of protein pairs exhibited a nominally significant correlation (*P <* 0.05) between predicted and observed expression/co-expression in all three models, but this percentage rose to over 99% in well-imputed pairs. We also observed a clear linear trend between the measured correlation of protein expression within well-imputed pairs and their imputed co-expression, further supporting the robustness of COWAS imputation models (Supplementary Figure 1). These results demonstrate that co-expression can be accurately imputed from genetic data, but further research beyond the scope of this paper is necessary to fully unravel the genetic architecture of protein co-expression.

It is known that *trans*-pQTLs are more likely than *cis*-pQTLs to exhibit horizontal pleiotropy, so using *trans*-pQTLs to predict expression may induce spurious associations between imputed expression and the outcome trait [9, 51, 52]. Since *trans*-pQTLs did not noticeably improve the imputation performance of co-expression imputation models and may confound protein-outcome associations, we decided to only use *cis*-pQTLs in our main analyses. Moreover, we chose to use lasso regression in our main analyses due to its computational efficiency and highest overall imputation quality. Model performance metrics for every combination of protein pair and model type are provided in Supplementary Data 2-13.

### COWAS identifies co-expressed proteins associated with complex traits

Having shown that COWAS is able to accurately impute both single-protein expression and protein co-expression, we applied it to three complex trait outcomes: low-density lipoprotein (LDL) cholesterol, Alzheimer’s disease (AD), and Parkinson’s disease (PD). We first considered LDL cholesterol because it is a well-studied quantitative trait with publicly available summary statistics from a very large GWAS meta-analysis. After demonstrating that COWAS can outperform standard PWAS for LDL cholesterol, we then applied our method to AD and PD, which are our primary disease areas of interest. For each outcome trait, we downloaded summary-level data from the largest publicly available GWAS study of European ancestry participants [42, 43, 45]. To ensure complete overlap between the genetic variants included in the imputation models and in the GWAS data, we re-trained COWAS models for each trait using only the intersection of variants found in both the UKB genotype data and in the trait’s GWAS. We also re-assessed the out-of-sample predictive performance of each model separately for each trait and only kept pairs with sufficiently high imputation accuracy, ensuring that differences between the GWAS datasets do not negatively impact the validity of association testing. As a result, the numbers of considered protein pairs somewhat differed among the three traits. For LDL cholesterol 613 pairs were accurately imputed, for AD there were only 564 well-imputed pairs, and for PD we retained 592 well-imputed pairs.

To compare our new approach with currently available methods, we performed a standard PWAS analysis for each protein included in the COWAS analyses. The same training samples, model types, and variant screening strategies were applied for both COWAS and PWAS. Namely, we selected the top 100 pQTLs for each protein in terms of their association effect sizes and used them as features in linear regression models with a lasso penalty. We also used the same LD reference panel derived from UKB data when computing effect sizes in both COWAS and PWAS. Full imputation performance metrics for all analyzed proteins and outcome traits are provided in Supplementary Data 14-20.

To account for multiple testing in COWAS, we performed a Bonferroni correction for the number of well-imputed protein pairs. Similarly, in standard PWAS we performed a Bonferroni correction for the number of well-imputed proteins. The primary purpose of our analyses is to compare the results of COWAS with those of PWAS on the same set of proteins, so correcting for the number of tests performed by each method is appropriate. In other contexts, a different multiple testing correction might be necessary. For example, one may decide to correct for three times the number of well-imputed pairs when simultaneously considering the COWAS interaction test and COWAS single-protein tests.

Our results demonstrate that COWAS is able to identify PPIs that have a significant genetically regulated effect on complex traits. For LDL cholesterol, which had the best-powered GWAS of the three traits we considered, we identified 38 protein pairs with a significant co-expression effect (Supplementary Figure 2). Of these protein pairs, 24 had at least one protein that was also identified by a standard PWAS analysis, while the rest were uniquely identified by our method. We also performed the COWAS global test on each pair to assess whether it has an overall effect on LDL cholesterol, which yielded 116 significant pairs. As expected, nearly all of those pairs contained at least one protein that was also detected by PWAS. However, our global test did identify 12 pairs with a significant effect on LDL cholesterol in which neither protein was significant when considered on its own, and five of those pairs did not even have a significant interaction term. This suggests that explicitly modeling co-expression can boost power relative to standard marginal tests, even when there is no statistically significant effect of co-expression on the outcome trait. Full results for LDL cholesterol, including the estimated effect sizes and standard errors for each pair, are provided in Supplementary Data 14.

Among protein pairs identified as significant for LDL cholesterol by either the COWAS global or interaction tests, 6% (7/123) were coded by genes located on the same chromosome, with an average distance of 22,961,613 base pairs (bp) between genes in those seven pairs. Among well-imputed protein pairs that were not identified as significant for LDL cholesterol, 9% (43/490) were coded by genes located on the same chromosome, with an average distance of 34,856,442 bp between genes in those 43 pairs. We also used the UKB protein expression data to calculate the average Pearson correlation between proteins in these sets of pairs, after normalizing and adjusting the expression data in the same way as done when training imputation models. On average, protein pairs identified as significant for LDL cholesterol by either the COWAS global or interaction tests had an absolute value correlation of 27.81%, while well-imputed but not significant protein pairs had a slightly higher average absolute value correlation of 31.72%.

Next, we explored whether the effects of genetically regulated co-expression on LDL cholesterol coincide with the interaction effects of observed expression on LDL cholesterol. We obtained individual-level data for LDL cholesterol from phase 2 of the UKB metabolomics study conducted by Nightingale Health [53] and applied the same data processing steps that we used for protein expression. Then we fit the model *Y* ∼ *A*+*B*+*A*∗*B* for every well-imputed protein pair, where *Y* denotes LDL cholesterol and *A, B* denote observed protein expression levels. Out of the protein pairs identified by the COWAS interaction test, we found that 74% (28/38) also had a nominally significant interaction term in this model (*P <* 0.05). These results suggest that the effects of observed and imputed co-expression on an outcome trait often align, even though they estimate different quantities from a statistical perspective. Full results from our validation study for LDL cholesterol are provided in Supplementary Data 15.

COWAS can also help to disentangle the effects of interacting groups of proteins on complex traits, thereby providing a richer picture of the functional consequences of genetically regulated molecular phenotypes. By visualizing significant COWAS pairs as a network, we identified complexes of mutually interacting proteins that modulate LDL cholesterol levels (Supplementary Figure 3). Moreover, our method can reveal when the effect of genetically regulated co-expression on a complex trait is in the opposite direction relative to the effects of the interacting proteins themselves. For example, we found that APOE and PLTP both decrease LDL cholesterol levels, while their co-expression is associated with increased LDL cholesterol levels (Supplementary Figure 4). In other cases, the direct and interaction effects may all be in the same direction, such as observed for the effects of APOE and AGRN on LDL cholesterol. We investigated whether there is a relationship between the directions of single-protein and co-expression effects on LDL cholesterol, but we did not find any significant trend (two-sided Fisher’s exact test *P* = 0.72).

### COWAS boosts power and corroborates known PPIs driving Alzheimer’s disease risk

We identified fewer significant protein pairs for AD compared to LDL cholesterol, but this was expected due to the lower power of the corresponding GWAS study. Yet here again, both the COWAS global test and the COWAS interaction test were able to detect significant protein pairs missed by standard PWAS (Figure 3a). Our method can also provide additional insights even in pairs where one protein was identified by PWAS. For example, we discovered five proteins that jointly affect AD risk together with CD33 (Figure 3b). Although CD33 itself was identified by standard PWAS, our application of COWAS revealed a fuller picture of the molecular pathways through which it affects AD.

**Fig. 3.**
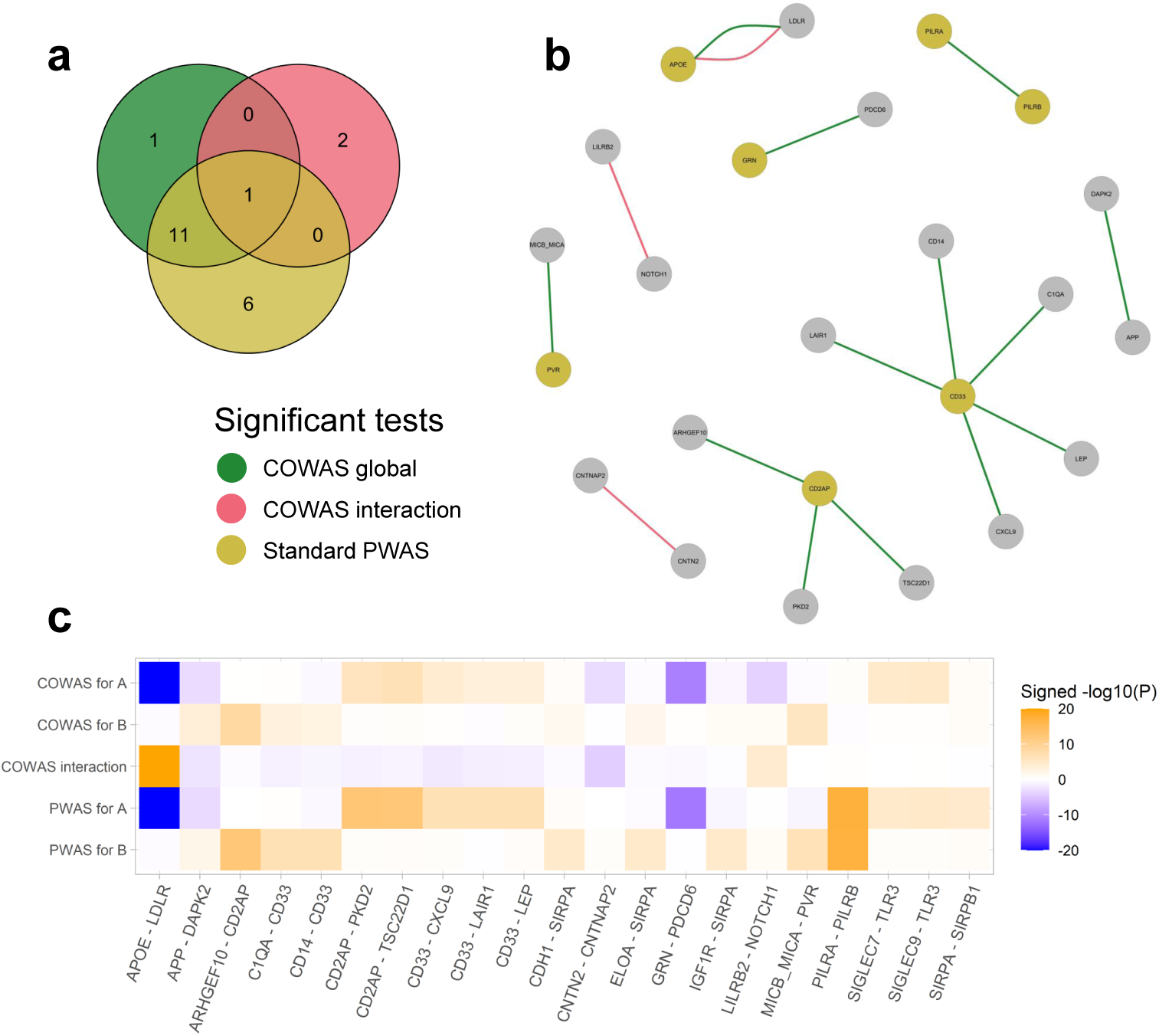
COWAS and PWAS results for Alzheimer’s disease. **a**, A Venn diagram displaying the numbers of protein pairs identified as significant for AD by the COWAS global test (green), the COWAS interaction test (pink), or a standard PWAS analysis (yellow). Here “standard PWAS” refers to pairs in which at least one of the proteins was identified by PWAS. **b**, A network diagram showing all of the protein pairs identified as significant for AD by either the COWAS global test (green edges) or the COWAS interaction test (pink edges). Node colors indicate whether each protein was identified as significant for AD by standard PWAS (yellow) or not (gray). **c**, A heat map displaying COWAS single-protein and interaction test results, as well as standard PWAS results, for all protein pairs identified as significant for AD by at least one of the three tests shown in **a**. Orange denotes positive effects and blue denotes negative effects, with color intensity corresponding to the *−* log_10_(*P*) value of the test. A and B refer to the first and second proteins listed in each pair, respectively. To facilitate visualization, the *−* log_10_(*P*) values were capped at 20.

Notably, the COWAS global test identified the pair comprised of amyloid-beta precursor protein (APP) and death-associated protein kinase 2 (DAPK2) as significant for AD (*P* = 1.25e-05), while a standard PWAS analysis failed to identify either of these proteins (*P* = 7.72e-04 for APP and *P* = 1.31e-02 for DAPK2). APP is concentrated in the synapses of neurons and is the precursor molecule for the generation of amyloid beta (Aβ), which contributes to the formation of amyloid plaques—a hallmark pathology in AD [54–56]. Yet despite the central role of APP in Alzheimer’s pathogenesis, standard PWAS lacked the power to identify it in our dataset. On the other hand, COWAS was able to boost power and attain statistical significance by jointly considering APP and a member of the DAPK family, which has also been previously implicated in late-onset AD [57].

Furthermore, COWAS discovered a highly significant effect of the interaction between APOE and LDLR on AD risk (*P <* 1e-50). Although APOE was also highly significant according to a standard PWAS analysis (*P <* 1e-50), LDLR was not (*P* = 0.48). This result is noteworthy, because LDLR was recently shown to be a receptor for APOE that preferentially binds lipidated APOE particles and plays an important role in Aβ clearance [58, 59]. Our results are consistent with this mechanistic explanation, since we found APOE and its interaction with LDLR to have opposite effects on AD (Figure 3c). Thus, COWAS provides strong support to the hypothesis that APOE and LDLR have a synergistic effect in Alzheimer’s pathogenesis, even after accounting for the direct effect of APOE on AD risk.

The other two significant interactions implicated by COWAS for AD are also likely true positives, further confirming the sensitivity and power of our approach. We identified a significant effect of co-expression between LILRB2 and NOTCH1 on AD (*P* = 5.73e-05), whereas standard PWAS failed to identify either protein (*P* = 0.80 for LILRB2 and *P* = 0.13 for NOTCH1). LILRB2 is a neuronal cell surface receptor that interacts with Aβ and is being studied as a promising therapeutic target for AD [60, 61], while NOTCH1 has been found to be differentially expressed in Alzheimer’s patients [62] and is potentially involved in neurodegeneration-related cell signaling disruptions [63]. Finally, the COWAS interaction test also discovered a significant effect of co-expression between CNTN2 and CNTNAP2 on AD (*P* = 8.42e-05), whereas standard PWAS again failed to detect either protein as significant (*P* = 0.94 and *P* = 0.38, respectively). The mechanisms by which these proteins are involved in Alzheimer’s pathology have not yet been thoroughly studied, but earlier genetic and functional genomic evidence indicates that they might play a role [64]. Full results for AD, including the estimated effect sizes and standard errors for each pair, are provided in Supplementary Data 16. Effect sizes, standard errors, and *P* values for the pairs shown in Figure 3 are also summarized in Supplementary Table 1.

Among protein pairs identified as significant for AD by either the COWAS global or interaction tests, 20% (3/15) were coded by genes located on the same chromosome, with an average distance of 12,452,978 bp between genes in those three pairs. Among well-imputed pairs that COWAS did not find to be significant for AD, a smaller proportion of 9% (47/549) were coded by genes located on the same chromosome, with a greater average distance of 38,351,197 bp between those 47 genes. Observed Pearson correlations between proteins in these two sets of pairs were similar, but non-significant pairs had a slightly higher average absolute value correlation of 31.65% compared to an average absolute value correlation of 28.72% among proteins in significant pairs.

To validate the robustness of the associations detected by COWAS, we repeated our analysis using the largest publicly available GWAS of clinically diagnosed lateonset AD. Unlike the European Alzheimer & Dementia Biobank (EADB) consortium GWAS used for our main results [43], the International Genomics of Alzheimer’s Project (IGAP) GWAS used for our validation study did not include proxy cases based on family history of dementia [44]. We repeated the model training process from scratch for variants present in the IGAP GWAS and obtained 601 well-imputed protein pairs. Of the protein pairs identified as significant by the COWAS global test with EADB GWAS data, 62% (8/13) also reached Bonferroni significance for the COWAS global test when using IGAP GWAS data. Although the three pairs identified by the COWAS interaction test with EADB GWAS data either failed quality control or did not reach Bonferroni significance with IGAP GWAS data, the COWAS interaction test did identify 10 other proteins using IGAP GWAS data whose co-expression with APOE significantly affects AD risk, reinforcing the same molecular pathway implicated in our EADB-based analysis. Standard PWAS discovered APOE, CD2AP, CD33, and PVR with both GWAS datasets, while LRRC25 was uniquely identified using the IGAP GWAS and GRN, PILRA, PILRB, SIGLEC7, SIGLEC9, and SIRPA were uniquely identified using the EADB GWAS. Results for protein pairs identified as significant using at least one of these three tests with IGAP GWAS data are shown in Supplementary Figure 5 and Supplementary Table 2. Full results for all protein pairs passing quality control are provided in Supplementary Data 17.

Finally, we considered an alternative, product-based formulation of COWAS (see Supplementary Note 1 for details). Instead of predicting the interaction of expression residuals, as done in the main version of our method, the product-based version of COWAS predicts the interaction of observed expression levels. We trained such product-based COWAS models on variants present in the EADB GWAS of AD and found 561 protein pairs to be well-imputed. The mean out-of-sample *R*^2^ for product-based co-expression imputation models was slightly higher than for any of the residual-based co-expression imputation models, and over 99% of well-imputed pairs again had a nominally significant correlation between imputed and observed expression (Supplementary Figure 6 and Supplementary Data 1). Using the interaction test in the product-based version of COWAS, we identified seven significant protein pairs: APOE and KLK10 (*P <* 1e-50), APOE and LCAT (*P <* 1e-50), APOE and LDLR (*P <* 1e-50), APOE and NOS3 (*P <* 1e-50), APOE and PLTP (*P* = 6.99e-22), LGALS9 and PVR (*P* = 1.27e-07), and LILRA3 and LILRB2 (*P* = 2.86e-07). Among these significant results, only the pair made up of APOE and LDLR was also identified by the interaction test in residual-based COWAS when using EADB summary statistics. Interestingly, however, three of these pairs were identified by the interaction test in residual-based COWAS when using IGAP summary statistics: APOE and LCAT, APOE and NOS3, and APOE and PLTP.

We also discovered 22 pairs with an overall effect on AD using the product-based COWAS global test and EADB summary statistics (Supplementary Figure 7 and Supplementary Table 3). Five of these pairs were also identified by the global test in residual-based COWAS using EADB summary statistics, and eight of them were identified by the global test in residual-based COWAS using IGAP summary statistics. Moreover, 77% (17/22) of these pairs contain at least one protein that was identified by the residual-based COWAS global test through a different interaction. For example, the product-based version of our method identified a significant overall effect of LGALS9 and PVR on AD (*P* = 9.50e-12). This pair does not appear in our main results, but the residual-based COWAS global test did identify PVR through its interaction with MICB MICA using EADB summary statistics, as well as through its interactions with CEACAM21 and PAEP using IGAP summary statistics. The other protein in that pair (LGALS9) belongs to the same family of galectin proteins as LGALS1, which was identified by the residual-based version of COWAS in a pair with APOE using IGAP summary statistics. These results suggest that both versions of COWAS largely pick up the same signals, but each can also provide complementary information. Full results for AD using the product-based version of COWAS are provided in Supplementary Figure 7, Supplementary Table 3, and Supplementary Data 18-19.

Among protein pairs identified as significant for AD by either the global or interaction tests in the product-based version of COWAS, 36% (8/22) were coded by genes located on the same chromosome, with an average distance of 5,504,443 bp between genes in those eight pairs. Among well-imputed pairs that product-based COWAS did not find to be significant for AD, a smaller proportion of 12% (66/539) were coded by genes located on the same chromosome, with a greater average distance of 24,701,012 bp between those 66 genes. Unlike in our results for residual-based COWAS, the average absolute value correlations were nearly identical between pairs that were significant for AD according to product-based COWAS and those that were not. Proteins in pairs identified as significant for AD had an average absolute value correlation of 31.26%, while proteins in well-imputed but not significant pairs had an average absolute value correlation of 31.93%.

### Co-expression analysis identifies SNCA interactions in Parkinson’s disease pathogenesis

For PD the COWAS global test identified all of the protein pairs that were discovered by the COWAS interaction test or by a standard PWAS analysis (Figure 4a). In addition to those pairs, the COWAS global test also uniquely identified an effect of GRK5 and SNCA on PD (*P* = 4.81e-06). This pair was not significant according to the COWAS interaction test (*P* = 2.75e-04) or according to standard PWAS (*P* = 0.04 for GRK5 and *P* = 1.65e-03 for SNCA). However, note that both of these proteins have been previously implicated in PD pathogenesis. Alpha-synuclein (SNCA) regulates the release of neurotransmitters from the axon terminals of presynaptic neurons, and insoluble forms of SNCA accumulate in the form of Lewy bodies, leading to nerve cell death and the development of PD symptoms [65–67]. As for GRK5, some evidence suggests that it plays a role in the pathogenesis of sporadic forms of PD [68]. These results further highlight the ability of COWAS to boost power relative to marginal approaches such as PWAS.

**Fig. 4.**
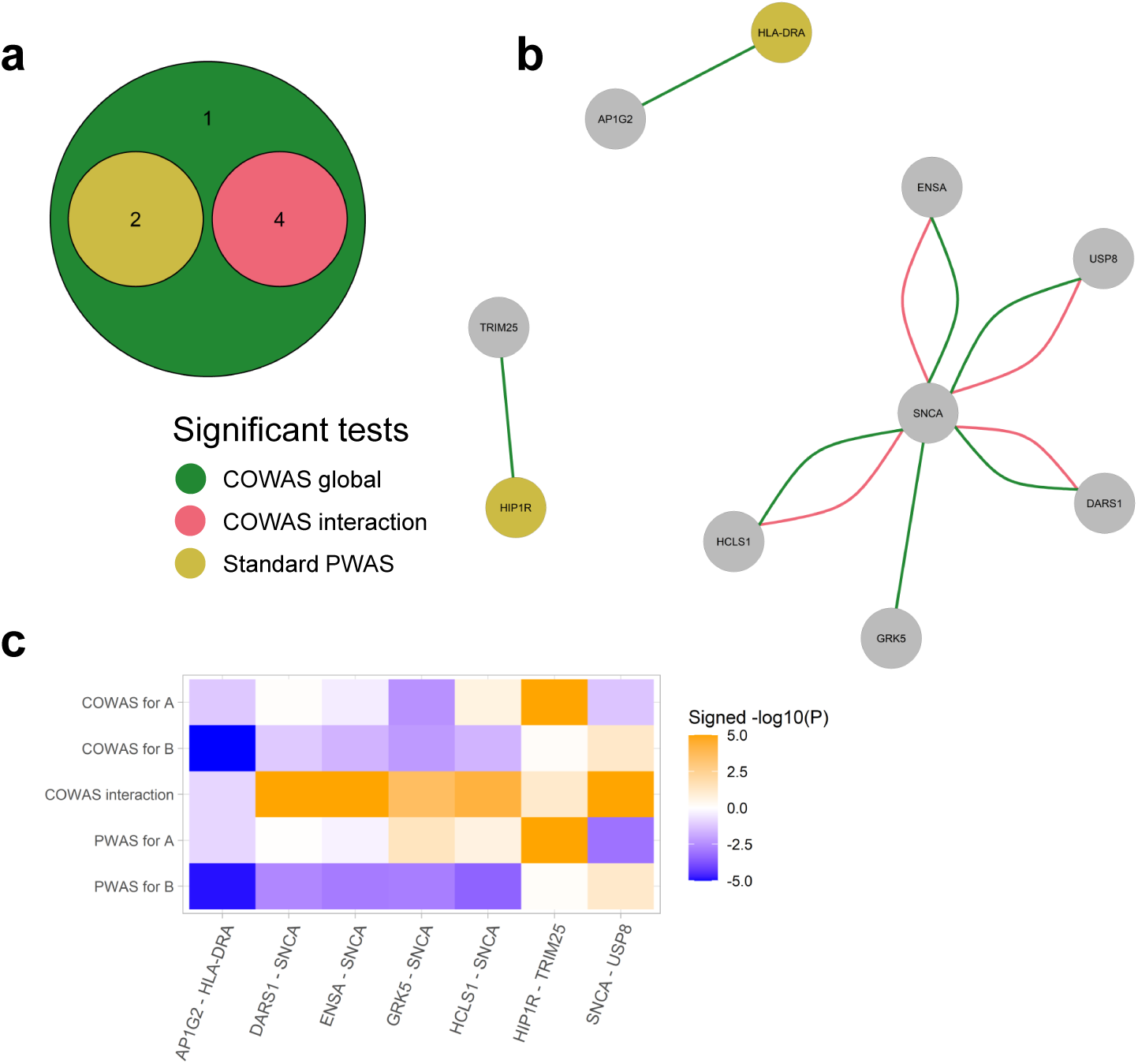
COWAS and PWAS results for Parkinson’s disease. **a**, A Venn diagram displaying the numbers of protein pairs identified as significant for PD by the COWAS global test (green), the COWAS interaction test (pink), or a standard PWAS analysis (yellow). Here “standard PWAS” refers to pairs in which at least one of the proteins was identified by PWAS. **b**, A network diagram showing all of the protein pairs identified as significant for PD by either the COWAS global test (green edges) or the COWAS interaction test (pink edges). Node colors indicate whether each protein was identified as significant for PD by standard PWAS (yellow) or not (gray). **c**, A heat map displaying COWAS single-protein and interaction test results, as well as standard PWAS results, for all protein pairs identified as significant for PD by at least one of the three tests shown in **a**. Orange denotes positive effects and blue denotes negative effects, with color intensity corresponding to the *−* log_10_(*P*) value of the test. A and B refer to the first and second proteins listed in each pair, respectively. To facilitate visualization, the *−* log_10_(*P*) values were capped at 5.

Interestingly, all four of the significant co-expression effects on Parkinson’s risk that were identified by COWAS are comprised of SNCA interacting with some other protein (Figure 4b). In particular, the COWAS interaction test identified significant effects on PD from genetically regulated co-expression between SNCA and DARS1 (*P* = 1.21e-13), SNCA and ENSA (*P* = 1.03e-08), SNCA and HCLS1 (*P* = 5.50e-05), and SNCA and USP8 (*P* = 3.64e-07). Note that co-expression between SNCA and each of these four proteins has a positive effect, even though the effect of SNCA itself is negative (Figure 4c). This suggests that a genetically regulated escalation of coexpression between SNCA and each of these proteins counteracts the effect of SNCA itself, illustrating potential avenues for therapeutic intervention.

Among the four proteins whose interactions with SNCA had an effect on PD, none were significant according to a standard PWAS analysis, with marginal PWAS *P* values ranging from *P* = 0.88 to *P* = 0.07. However, the COWAS discoveries are reasonable in light of previous research. For example, USP8 is a deubiquitinase that has also been found in Lewy bodies and plays a role in determining SNCA levels [69, 70]. ENSA has been shown to interfere with SNCA self-assembly and thereby alleviate its neurotoxicity [71], and variants in HCLS1 binding protein 3 were found to be associated with the related condition of essential tremor (but not PD itself) [72]. We are not aware of any existing evidence for the role of DARS1 in Parkinson’s pathogenesis, but its identification by COWAS points to a potential avenue for further research. Full results for PD, including the estimated effect sizes and standard errors for each pair, are provided in Supplementary Data 20. Effect sizes, standard errors, and *P* values for the pairs shown in Figure 4 are also summarized in Supplementary Table 4.

Finally, we again compared the physical characteristics of significant protein pairs and those that were well-imputed but not significant. Among the seven protein pairs that were identified as significant for PD by either the COWAS global test or the COWAS interaction test, none were made up of proteins coded by genes located on the same chromosome. On the other hand, among pairs that were well-imputed but not identified as significant for PD by COWAS, 9% (52/585) were made up of proteins coded by genes located on the same chromosome. Genes coding for the proteins in those 52 pairs had an average pairwise distance of 31,453,275 bp between them. Unlike in our results for LDL cholesterol and AD, the average absolute value correlation between proteins was much higher in pairs identified as significant for PD (47.02%) than in well-imputed but non-significant pairs (31.85%).

## Discussion

In this paper we introduced the co-expression-wide association study (COWAS) method, which is the first statistical framework for identifying gene or protein pairs whose genetically regulated co-expression is associated with complex traits. COWAS extends the two-stage least squares approach underlying TWAS/PWAS by explicitly imputing the interaction of expression residuals for pairs of exposures, which we interpret as a proxy for genetically regulated gene–gene or protein–protein interactions. This enables COWAS to jointly test for the effects of genetically regulated expression and co-expression on a complex trait of interest, thereby boosting power relative to existing methods and helping to disentangle the functional mechanisms by which molecular exposures influence the outcome trait. We also introduced an alternative version of COWAS that imputes the product of normalized expression levels instead of the product of their residuals, which more closely resembles a standard interaction model. Both versions can successfully identify disease-relevant interacting protein pairs, but they differ in terms of interpretation. Regardless of the type of model chosen, association testing in COWAS can be performed using summary statistics, making it easy to apply our method to any trait for which GWAS summary data are available.

In our application of COWAS to the UKB Pharma Proteomics Project dataset, we first explored the performance of different regression models in imputing genetically regulated co-expression. Then we used our method to identify protein pairs associated with three complex traits. COWAS was able to discover biologically relevant co-expressed proteins associated with each of the three traits, highlighting the importance of interaction effects in driving complex disease risk. Notably, COWAS identified a number of protein pairs with a significant interaction term or a significant overall effect in which neither protein was significant when tested independently via standard PWAS. These results underscore the importance of considering interaction effects and demonstrate that standard TWAS/PWAS approaches may be missing important sources of signal. Moreover, our results illustrate how COWAS can be used to implicate groups of proteins in complex disease risk and to distinguish between direct and interaction effects, providing a more complete picture of the molecular pathways that mediate genetic risk on downstream traits. In future work it will be important to explore the performance of residual-based COWAS and product-based COWAS in disease areas other than neurodegeneration, and to assess the relative importance of co-expression across different traits.

Notwithstanding the many advantages of COWAS, our approach has several limitations. First of all, COWAS only considers one pair of molecular units at a time. Although these pairwise results can be visualized as a network, COWAS does not simultaneously model the genetic regulation of entire protein complexes. Proteins may interact in larger, multi-protein interaction networks with nontrivial topological structures [73–75], and it is possible that genetic variation may impact higher-order network properties. Extending COWAS to allow for other types of interactions among more than two exposures at a time could illuminate additional disease-relevant genes and proteins, but it is not obvious how to do so in a computationally efficient way. Furthermore, we found that the predictive capacity of protein co-expression imputation models is lower than that of expression imputation models for individual proteins. This was expected given the difficulty of ascertaining interaction effects in general, yet even so we were able to obtain sufficiently good imputation quality for over a thousand protein pairs. However, more work should be done to explore different machine learning algorithms for training co-expression imputation models. In this study we also did not focus on identifying coQTLs or investigating their biological properties. Past research has shown that genetic variation can alter co-expression relationships [36–39, 46–48] and we have now demonstrated that models can be trained to accurately predict co-expression from genotype data, but more work is necessary to understand the genetic architecture of co-expression and to quantify the extent to which coQTLs overlap with quadratic QTLs and variance QTLs.

Because COWAS is based on the statistical framework of instrumental variable (IV) regression, it also inherits many of the fundamental limitations of TWAS and other IV regression methods. In particular, association results from TWAS and related methods may be confounded due to the presence of horizontal pleiotropy [23, 51, 76–78]. If the genetic variants used to impute expression have a causal effect on the outcome trait through some exposure that is not included in the IV regression model, effect estimates will be biased. Our method partially mitigates this issue by including two related exposures in each model, but beyond that COWAS is not robust to violations of the IV assumptions necessary for causal inference [30, 76, 79]. Furthermore, sample overlap between the datasets used for model training and association testing is known to bias estimates of the exposure-outcome relationship obtained via IV regression [80]. Our decision to maximize power by using the largest available GWAS for each outcome trait may have led to some overlap between the individuals present in the GWAS cohorts and the individuals included in our training dataset. Lastly, transcriptome and proteome imputation models are not portable across human tissues and ancestry groups [11, 13, 17, 24, 26, 81–86]. In this study we subset the UKB data to the largest genetically-inferred ancestry subgroup, which roughly corresponds to White British individuals, and correspondingly used GWAS studies conducted on European individuals for our three outcome traits. An extension of COWAS to handle multiple tissue types and multiple ancestry groups would expand the diversity and relevance of its applications.

Historically, the field of human genetics has exclusively focused on studying linear and marginal effects. This is exemplified by the popularity of GWAS and TWAS/PWAS analyses, which only consider one genetic variant or one functional molecular unit at a time. By providing a simple yet powerful approach for analyzing genetically regulated gene or protein co-expression using existing biobank data, our work joins the growing body of evidence emphasizing the limitations of that historical paradigm. The COWAS method exhibits high statistical power, provides flexibility in modeling direct and interaction effects, and is easy to use. We envision that COWAS, along with its future improvements and extensions, will enhance the interpretation of genomic findings and lead to the discovery of new biological insights and therapeutic targets.

## Methods

### Modeling genetically regulated co-expression

The COWAS method is applied to one outcome trait and two molecular exposures at a time. Let *A_i_* and *B_i_* be real-valued random variables that represent the expression or abundance levels of the two exposures for some individual *i*. Further, let ***Z****_A,i_* be a random vector of length *p_A_*whose entries contain dosage values in individual *i* for the *p_A_*xQTLs that regulate the first exposure. Similarly, let ***Z****_B,i_* be a random vector of length *p_B_* containing dosage values in individual *i* for the *p_B_* xQTLs that regulate the second exposure. Finally, let *Y_i_* be a real-valued random variable representing the value of the outcome trait in individual *i*.

Just like in standard TWAS and PWAS, we assume that the genetic component of the expression of each molecular exposure can be modeled as a linear combination of its xQTL genotypes. That is, we assume the models

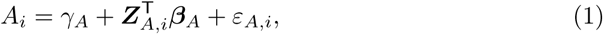

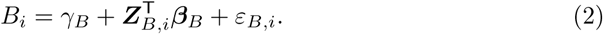

Here ***β****_A_* ∈ 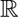*^pA^* and ***β****_B_* ∈ 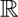*^pB^* are the true xQTL weights, while *γ_A_, γ_B_* ∈ 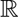 are the true intercepts. The error terms *ε_A,i_* and *ε_B,i_* are real-valued random variables with mean zero.

What sets COWAS apart from previous methods, however, is that we also model the genetically regulated co-expression of the two functional units instead of analyzing them independently of each other. Doing so requires careful consideration of what is meant by the genetic regulation of co-expression. To our knowledge, we are the first to introduce a formal statistical definition of this concept. Let

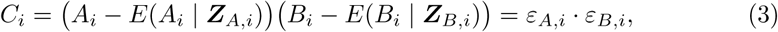

where in practice the error terms are replaced by residuals from fitting Equation (1) and Equation (2). We define the genetically regulated co-expression of the two exposures to be *E*(*C_i_* | ***Z****_A,i_,* ***Z****_B,i_*). Note that the quantity *C_i_*is an interaction between expression residuals, after removing the main effects of genetic variation on expression. Thus, our definition of genetically regulated co-expression can be thought of as the genetic component of this interaction term.

We assume that the genetically regulated co-expression of a pair of molecular phenotypes can be modeled as a linear combination of their xQTL genotypes. That is, we assume the model

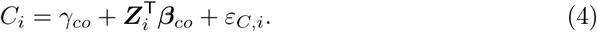

Here ***Z****_i_* is a random vector of length *p* whose entries contain dosage values in individual *i* for all *p* xQTLs included in ***Z****_A,i_* and ***Z****_B,i_*, with *p* being the number of unique variants in the union of xQTLs for the two exposures. (If there is no overlap among the xQTLs for the two exposures, then *p* = *p_A_* + *p_B_* and ***Z****_i_* is formed by simply stacking ***Z****_A,i_* and ***Z****_B,i_* on top of each other.) The parameter ***β****_co_* ∈ 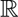*^p^* is the true vector of weights for the genetic variants in ***Z****_i_*, the term *γ_co_* ∈ 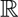 is the true intercept, and the error term *ε_C,i_*is assumed to be a real-valued random variable with mean zero.

We argue that *E*(*C_i_* | ***Z****_i_*) is an apt statistical representation of the biological concept of genetically regulated co-expression. In general, correlation between the expression levels of two molecular phenotypes can be attributed to some combination of the following three factors:

1. Genetic effects on mean expression levels. That is, genetic variation can induce correlation between *E*(*A_i_* | ***Z****_i_*) and *E*(*B_i_* | ***Z****_i_*) across individuals. This may cause the expression levels themselves to be correlated.
2. Genetic effects that modulate the amount of correlation between the two phenotypes. That is, the correlation between *ε_A,i_* and *ε_B,i_* across individuals can change with respect to genetic variation. This may likewise cause the expression levels themselves to be correlated.
3. Factors unrelated to genetics. For example, a shared tissue environment or various other environmental effects may cause expression levels to be correlated.

The concept of “genetically regulated co-expression” should naturally correspond to the second factor: genetic regulatory effects on the correlation between expression levels of the two phenotypes. Notice that our formulation of *C_i_* explicitly removes the first factor by subtracting out the linear genetic component of each exposure’s expression. Moreover, the effects of any factors unrelated to genetics should be constant with respect to genetic variation, so environmental effects on mean expression levels will be mostly absorbed by the intercept terms *γ_A_* and *γ_B_*, while environmental effects on *C_i_*will be mostly absorbed by *γ_co_*. As a result, ***β****_co_* will largely avoid reflecting the third factor. Therefore, ***β****_co_* has the desired interpretation of representing the effect of genetic variation on co-expression without being tainted by marginal genetic effects or environmental effects.

Importantly, our definition of genetically regulated co-expression in terms of the expected product of expression residuals is not equivalent to a simple interaction term between the two exposures’ observed or imputed expression levels. In Supplementary Note 1 we define an alternative, product-based version of COWAS in which *C_i_* is replaced with *T_i_*= *A_i_* − *E*(*A_i_*) *B_i_* − *E*(*B_i_*). Both versions can successfully identify disease-relevant protein pairs, but our primary formulation provides a more elegant interpretation because it only models genetic effects that directly modulate the amount of correlation between exposures. The residual-based and product-based models are both available in our R implementation of COWAS (https://github.com/mykmal/cowas).

In the second stage of COWAS, we model the effect of genetically regulated co-expression on the outcome trait while also accounting for direct effects from *E*(*A_i_* | ***Z****_A,i_*) and *E*(*B_i_* | ***Z****_B,i_*). We assume that the outcome trait depends on a linear combination of the genetically regulated expression levels of both molecular exposures as well as their genetically regulated co-expression. Therefore, our model for the outcome trait is

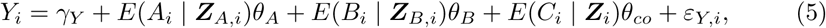

where *θ_A_, θ_B_, θ_co_* ∈ 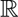 are the true effect sizes, γ*_Y_* ∈ 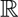 is the true intercept, and the error term *ε_Y,i_*is a real-valued random variable with mean zero. Each coefficient in our stage 2 model has a straightforward interpretation. The coefficient *θ_A_*is the effect of the genetic component of the first exposure on the outcome trait, after accounting for the genetic component of the other exposure and for the two exposures’ genetically regulated co-expression. Similarly, *θ_B_* is the effect of the genetic component of the second exposure on the outcome trait, after accounting for the genetic component of the first exposure and for the two exposures’ genetically regulated co-expression. Finally, *θ_co_*is the effect of genetically regulated co-expression on the outcome trait, after accounting for the genetically regulated expression levels of both exposures.

### Two-sample model estimation and hypothesis testing

COWAS model estimation is based on the two-sample, two-stage least squares regression framework, akin to standard TWAS and PWAS. Suppose we have two individual-level datasets sampled from the same population with sample sizes *n*_1_ and *n*_2_. Let ***A***, ***B*** ∈ 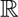*^n^*^1^ be observed vectors of the expression or abundance levels of the two exposures, as measured in the first dataset. Also let **Z**_A_ ∈ 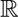*^n^*^1 ×^*^pA^* be the observed genotype matrix of *p_A_* xQTLs for the first exposure, and let **Z**_B_ ∈ 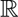*^n^*^1 ×^*^pB^* be the observed genotype matrix of *p_B_* xQTLs for the second exposure, as genotyped in the same dataset of *n*_1_ individuals. Then let **Z** ∈ 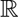*^n^*^1 ×^*^p^* be the observed joint genotype matrix of all *p* xQTLs. Analogously we define 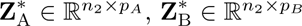, and **Z**^∗^ ∈ 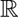*^n^*^2 ×^*^p^* for the second dataset. Finally, let ***Y*** ^∗^ ∈ 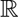*^n^*^2^ be an observed vector of outcome trait measurements for each individual in the second dataset. All of these vectors and each column of these matrices are assumed to be centered around zero and scaled to have a variance of one. Note that genotype data for all *p* xQTLs is available in both datasets, but the molecular exposures are only measured on the *n*_1_ samples in the first dataset, while the outcome trait is only measured on the *n*_2_ samples in the second dataset.

In the model training stage, ***A*** and ***B*** are regressed on **Z**_A_ and **Z**_B_, respectively, to obtain estimates of the parameter vectors ***β****_A_* and ***β****_B_*. We used penalized linear regression to obtain 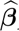*_A_* and 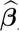*_B_*, but any other variable selection methods for linear regression can also be used. Then COWAS imputes expression for each exposure on that same dataset. That is, we obtain the predictions

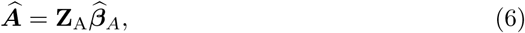

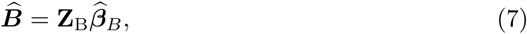

on the same *n*_1_ individuals used for model training. Next, COWAS computes the Hadamard product 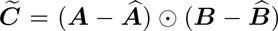. Finally, 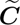 is regressed on **Z**, yielding the fitted coQTL weight vector 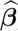*_co_*. Once again, we used penalized linear regression to estimate these variant weights.

In the association testing stage, the fitted weights 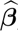*_A_*, 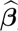*_B_*, and 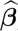*_co_* are used to impute expression and co-expression for the *n*_2_ samples in the second dataset. That is, we compute

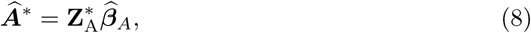

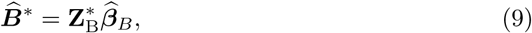

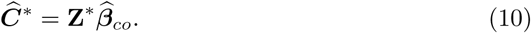

Lastly, COWAS fits a multiple linear regression model with ***Y*** ^∗^ as the outcome and 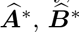 and 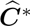 as predictors to obtain estimates of the parameters *θ_A_*, *θ_B_*, and *θ_co_*. Any linear model hypothesis tests can be performed on the estimated coefficients 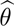*_A_*, 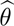*_B_*, and 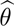*_co_*. In this paper, we primarily considered an interaction test and a global test.

### Interaction test

To determine if co-expression has an effect on the outcome trait, we test the hypothesis *H*_0_ : *θ_co_* = 0 against its two-sided alternative using a Wald test. Namely, the test statistic is 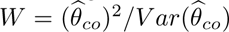, which asymptotically follows a *χ*^2^ distribution with one degree of freedom under *H*_0_.

### Global test

To determine if the two exposures have an overall effect on the outcome trait, we use an *F* test to check whether our second-stage model fits the data better than an intercept-only model. Namely, the test statistic is *F* = 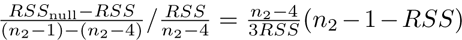, where *RSS* is the residual sum of squares from our fitted second-stage model, and *RSS*_null_ = *n*_2_ −1 is the residual sum of squares from an intercept-only model. This statistic follows an *F* distribution with (3*, n*_2_ − 4) degrees of freedom under the null hypothesis that the two models are equivalent. Note that as *n*_2_ → ∞ this *F* statistic will approach a *χ*^2^ distribution with three degrees of freedom under the null, so our global test can also be thought of as an approximation to a *χ*^2^ test.

The interaction test and the global test can help to disentangle the effects of coexpression from the direct effects of the individual exposures. For example, if the global test rejects its null hypothesis but the interaction test does not, we can conclude that the molecular exposures primarily influence the outcome trait through pathways not involving their co-expression. Our implementation of COWAS in R also provides *P* values for Wald tests on *θ_A_* and *θ_B_*, enabling users to test whether each exposure has a significant effect while accounting for the other exposure and for their co-expression. However, we do not focus on those tests in this paper because their interpretation is not as interesting from a biological perspective.

The model training stage of COWAS (stage 1) requires individual-level data, but the association testing stage (stage 2) can be performed using fitted imputation model weights and GWAS summary statistics for the outcome trait. In Supplementary Note 2 we derive a summary-level version of the second stage following the approach introduced in the multivariate imaging-wide association study (MV-IWAS) method [14, 16], which is related to other methods in the field of Mendelian randomization [87, 88]. We also provide full derivations for both individual-level and summary-level versions of standard TWAS/PWAS in Supplementary Note 3. All results reported in this paper were obtained using the versions of COWAS and PWAS based on summary data.

### Expression and co-expression imputation models

Any statistical or machine learning algorithms can be used to build expression and coexpression imputation models for COWAS when individual-level data are available for the outcome trait. In order to perform the testing stage of COWAS using summary-level GWAS data, however, the stage 1 models must be built using an algorithm that fits weights for a linear combination of SNPs. In this paper, we evaluated penalized linear regression models with three different penalties: the elastic net penalty, the lasso penalty, and the ridge penalty. All models were trained using the glmnet package in R. The *α* hyperparameter was set to *α* = 0.5 for elastic net regression, *α* = 1 for lasso regression, and *α* = 0 for ridge regression. The *λ* hyperparameter, which controls the strength of the penalty, was chosen through 10-fold cross validation. Our implementation of COWAS also provides the option for linear regression with stepwise variable selection, but we did not apply that method here due to its much longer runtime.

To decrease the runtime of COWAS and make it scalable to tens of thousands of protein pairs, we pre-screened genetic variants before including them as features in the penalized regression models. Namely, we utilized the approach known as Sure Independence Screening (SIS) [89]. SIS relies on the theoretical result that variable selection based on marginal correlations will consistently select important predictors in ultrahigh-dimensional models. Thus, important variants for predicting protein expression can be consistently selected by restricting the initial set of variants considered by each model to a moderate number of top pQTLs. This is a fairly common practice in TWAS and PWAS [16, 19, 20, 90]. We performed variant pre-screening by first conducting a proteome-wide pQTL mapping study to compute the association between each variant and the expression of each protein, after normalizing and adjusting for covariates. Then we considered two approaches for pre-screening predictive variants. In *P* value screening, we ranked variants by their pQTL *P* values and kept the top 100. In effect size screening, we instead ranked variants by the absolute values of their pQTL effect sizes and again kept the top 100. In both cases, the initial feature set for the co-expression model was taken to be the union of the top-ranked pQTLs for the two proteins. Note that for models trained with *cis*-pQTLs only, we selected the top 100 pQTLs located within the *cis* region of the gene that codes for each protein. Regardless of whether *P* values or effect sizes are used to rank variants, considering only strong pQTLs greatly decreases the time taken to train expression imputation models without significantly sacrificing prediction quality, making our method computationally feasible to apply on large-scale biobank datasets.

Following the convention in standard TWAS and PWAS, we ensured that only well-imputed protein pairs are considered in the testing stage. COWAS assesses the predictive performance of each model by calculating the correlation between imputed and measured expression on a held-out test set. In particular, we randomly selected 80% of the available samples for each protein pair to train imputation models and used the remaining 20% to test their predictive performance. For single-protein models, we calculated the correlation between imputed (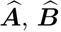) and measured (***A***, ***B***) expression on the test set. Recall that the outcome in the co-expression model, on the other hand, is a quantity estimated using measured expression levels as well as predictions from single-protein models. Thus, to evaluate the performance of the co-expression model, we first obtained 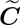 using co-expression and single-protein models trained only on the 80% training set. Then we calculated 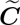 on the 20% test set, but this time using single-protein models trained on all available data. The out-of-sample correlation for the co-expression model was calculated as the correlation between these estimates of 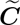 and 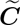 on the test set. After assessing predictive performance, all three models were re-trained on the full dataset to obtain the final xQTL weight vectors.

Only pairs in which all three models had out-of-sample correlations greater than 0.03 were used for hypothesis testing. We decided to filter out low-quality imputation models based on their correlations instead of their coefficients of determination (*R*^2^) to make sure that we do not select any models whose predictions are negatively correlated with observed expression. Although our threshold is somewhat less stringent than the *R*^2^ ≥ 0.01 and *R*^2^ ≥ 0.005 thresholds commonly used in PWAS [7, 15, 19, 27, 31, 84, 91], a lower threshold is appropriate given the difficulty of predicting interaction effects. Moreover, previous theoretical results and simulation studies have shown that noise or errors in an exposure variable will not inflate the type I error rate in association tests of the exposure-outcome relationship, as long as the errors are independent of the outcome [92, 93]. The only downside of using lower-quality imputation models for association testing is lower power, but this can be partially counteracted by using a sufficiently well-powered GWAS for the outcome trait [81, 94].

### Data processing and quality control

We trained protein expression and co-expression imputation models on individual-level data from the UKB. First we downloaded genotype data for 92,457,702 autosomal markers and 487,363 samples. Imputation and extensive quality control checks had already been performed on the provided data as detailed previously [40]. We then subset the data to only keep high-quality samples of genetically-inferred White British ancestry with no relatives of third degree or closer, using indicators provided by the UKB. Furthermore, we subset the data to individuals who have proteomic measurements available at the baseline visit. After these steps, 36,171 samples remained.

We also performed additional variant-level quality control on the UKB genotype data. In particular, we removed all variants with a missingness rate greater than 10% across the remaining individuals, those with a minor allele count (MAC) less than 100, those with a minor allele frequency (MAF) less than 1%, and those that failed a Hardy–Weinberg equilibrium test with *P <* 10e-15. To facilitate matching up variants between UKB data and outcome trait GWAS data, we also removed all variants lacking an rsID and those that are palindromic. Finally, we pruned the variants to *r*^2^ *<* 0.8 with a 1,000 bp window and a step size of 100 bp. After these steps, 1,689,714 variants remained. All sample-level and variant-level quality control was done in PLINK 2.0 (https://www.cog-genomics.org/plink/2.0).

Next, we computed genetic principal components (PCs) from the quality-controlled genotype data. Before computing PCs, we applied a few more data processing steps in addition to those described in the previous paragraph. In particular, we additionally removed all variants in regions of long-range LD [40], and then we pruned the remaining variants to a strict threshold of *r*^2^ *<* 0.1 with a 1,000 bp window and a step size of 100 bp. The computation of genetic PCs was also done in PLINK 2.0.

Proteomic profiling in blood plasma was performed by the UKB Pharma Proteomics Project using the antibody-based Olink Explore 3072 proximity extension assay, which measured 2,941 protein analytes across eight panels and captured 2,923 unique proteins [41]. Various quality control checks and normalization had already been performed as described previously [41]. We downloaded Normalized Protein eXpression (NPX) values for 2,923 proteins in 53,073 samples. Then we subset the proteomic data to the set of 36,171 individuals described above and only kept protein abundance measurements from the baseline visit. The sample sizes for individual proteins ranged from 106 to 35,581 individuals, with a median sample size of 30,526 individuals. Rather than imputing missing values, we took the intersection of samples with non-missing data within each protein pair when training imputation models.

Next, we normalized the NPX values using a rank-based inverse normal transformation. In particular, we utilized the commonly-used Blom transform with an offset of 0.375 and ties broken by averaging. Following the transformation, we regressed out the following standardized covariates: age, age^2^, sex, age * sex, age^2^ * sex, UKB assessment center, genotyping array type, and the first 20 genetic PCs. The inverse-normal transformation and covariate adjustment were applied separately for each pair, using the samples with non-missing data for the two proteins in the pair. The protein expression levels obtained after normalizing and adjusting for covariates were used in all downstream analyses.

### Protein annotations

We trained models that include pQTLs screened from across the genome, as well as models that only include *cis*-pQTLs. To identify the *cis*-SNPs for each protein, we obtained start and end positions for the genes coding each assayed protein from annotations provided by the UKB Pharma Proteomics Project [41], and then lifted them over to the hg19 genome build using the UCSC LiftOver web tool (https://genome.ucsc.edu/cgi-bin/hgLiftOver). We defined the *cis* region for each gene as beginning 500,000 bp upstream of its transcription start site and ending 500,000 bp downstream of its transcription end site. For proteins coded by several genes, we only considered the first gene listed in the UKB annotation file.

Training imputation models for every possible pair of proteins would have been computationally infeasible. To reduce the number of models that need to be trained and tested, we decided to only train models for pairs of proteins with some prior evidence of PPIs. Since non-interacting proteins are less likely to be co-expressed, it is also less likely that COWAS will be able to successfully predict their co-expression, and such models would get filtered out by our imputation quality threshold. Thus, computational time can be greatly reduced without significantly sacrificing important signals by training models only for pairs with prior evidence of interactions. This is analogous to how the FUSION implementation of TWAS only attempts model training for heritable genes [8]. However, excluding some proteins or protein pairs before model training may introduce selection bias. This will happen when the filtering criteria are associated with the outcome, introducing an extraneous “flow” of influence between the exposures and the outcome [95]. To minimize this possibility, we only considered PPIs established using experimental evidence instead of computational methods, ensuring that no information on downstream phenotypes (e.g., disease status) was considered when identifying interactions.

In particular, we restricted model training to protein pairs listed in the Human Integrated Protein–Protein Interaction rEference (HIPPIE) database [50]. To identify those pairs, we downloaded version 2.3 of the HIPPIE database and mapped each protein in the database to its gene name using the UniProt ID Mapping web tool (https://www.uniprot.org/id-mapping). The gene names were then matched with protein annotations from the UKB, and protein pairs present in the HIPPIE database were retained. Note that we considered all protein pairs listed in the HIPPIE database, regardless of their interaction confidence score. This reduced the number of protein pairs from 4,270,503 to 28,286. Finally, we also excluded proteins coded by non-autosomal genes. This further reduced the total number of proteins to 2,833 and the total number of pairs to 26,433.

### GWAS data for outcome traits

We considered three complex traits as outcomes in our application of COWAS: lowdensity lipoprotein (LDL) cholesterol, Alzheimer’s disease (AD), and Parkinson’s disease (PD). The testing stage of COWAS was performed using summary-level data from the largest available GWAS study for each trait. Since none of the GWAS studies provided *Z* scores in their summary data, we computed them by dividing each variant’s effect size by its standard error.

For LDL cholesterol levels, we downloaded GWAS summary statistics data from the Global Lipids Genetics Consortium (GLGC) [42]. The GLGC aggregated GWAS results from 1,320,016 individuals of European ancestry across 146 cohorts. Their meta-analysis provided summary statistics for 47,006,483 genetic variants and five lipid traits, including LDL cholesterol. Although the authors also conducted a multi-ancestry meta-analysis, we used results that were meta-analyzed solely in the European cohorts to ensure consistency with the genetic ancestry of UKB participants. A total of 1,624,628 genetic variants remained after harmonization with our quality-controlled UKB genotype data.

For AD status, we downloaded GWAS summary statistics data from the European Alzheimer & Dementia Biobank (EADB) consortium [43]. Namely, we used their stage 1 GWAS of AD and related dementias in individuals of European ancestry. The stage 1 GWAS was a meta-analysis based on 39,106 clinically diagnosed cases, 46,828 proxy cases (with disease status inferred from parental history), and 401,577 controls. Summary statistics for 21,101,114 genetic variants were provided, of which 1,435,986 remained after harmonization with our quality-controlled UKB genotype data.

Although the EADB GWAS is the largest GWAS available for Alzheimer’s or dementia, some have questioned its quality because the EADB meta-analysis relied in part on proxy cases, where participants’ disease status was imputed from their family history of AD instead of being clinically diagnosed [96]. To validate our main results, we also considered the largest GWAS of AD that does not contain proxy cases: the International Genomics of Alzheimer’s Project (IGAP) consortium GWAS of late-onset AD [44]. In particular, we utilized results from the stage 1 IGAP metaanalysis of non-Hispanic Whites, which included 21,982 clinically diagnosed cases and 41,944 cognitively normal controls. Summary statistics for 11,480,632 genetic variants were provided, of which 1,374,921 variants remained after harmonization with our quality-controlled UKB genotype data.

For PD status, we downloaded GWAS summary statistics data from the International Parkinson Disease Genomics Consortium (IPDGC) [45]. The IPDGC GWAS is also a meta-analysis, aggregating associations across 17 cohorts with individuals of European ancestry. Their main analysis included 37,688 clinically diagnosed cases, 18,618 proxy cases (with disease status inferred from first-degree relatives), and 1,417,791 controls. However, the publicly available summary statistics exclude three studies with individuals from 23andMe due to data sharing restrictions. We used the publicly available GWAS data in our analysis, which was based on 15,056 clinically diagnosed cases, 18,618 proxy cases, and 449,056 controls. Summary statistics were provided for 17,443,094 genetic variants, of which 1,393,959 remained after harmonization with our quality-controlled UKB genotype data.

## Data availability

Genotype, covariate, and protein expression data from the UK Biobank are available through the UK Biobank data access process (https://www.ukbiobank.ac.uk/enable-your-research). Access to the UK Biobank data was approved through UK Biobank Application #35107. Annotations for proteins assayed by the UK Biobank Pharma Proteomics Project are publicly available on Synapse (https://www.synapse.org/Synapse:syn51364943). Protein pairs with known interactions are publicly accessible in the HIPPIE web tool (https://cbdm-01.zdv.uni-mainz.de/~mschaefer/hippie). Publicly available GWAS summary statistics for cholesterol levels were downloaded from the Global Lipids Genetics Consortium website (https://csg.sph.umich.edu/willer/public/glgc-lipids2021). Publicly available GWAS summary statistics for Alzheimer’s disease and Parkinson’s disease were obtained from the NHGRI-EBI GWAS Catalog (https://www.ebi.ac.uk/gwas) under accession numbers GCST90027158, GCST007511, and GCST009325.

## Code availability

Our software for COWAS is implemented in R and made available on GitHub under a GPL-3.0 open source license at https://github.com/mykmal/cowas. This GitHub repository also contains the scripts used for data quality control and batch processing. Fitted model weights for all protein expression and co-expression imputation models trained in this study are provided on Synapse at https://synapse.org/cowas.

## Acknowledgements

We thank the two anonymous reviewers who provided thoughtful and constructive feedback on earlier versions of this paper. This work was supported by the National Institutes of Health (NIH) under grants R01 AG065636 and RF1 AG067924. The content is solely the responsibility of the authors and does not necessarily represent the official views of the NIH. We also acknowledge the Minnesota Supercomputing Institute (MSI) at the University of Minnesota for providing high-performance computing resources.

## Author contributions

W.P. conceived and supervised the project. M.M. developed the method, implemented the software, and performed the analyses. M.M. drafted the manuscript and W.P. proofread it.

