## Supplementary Information for "Co-expression-wide association studies link genetically regulated interactions with complex traits"

Mykhaylo M. Malakhov[1] and Wei Pan[1,*]

[1]Division of Biostatistics and Health Data Science, School of Public Health, University of Minnesota, Minneapolis, MN, USA.

#### 1 Supplementary figures

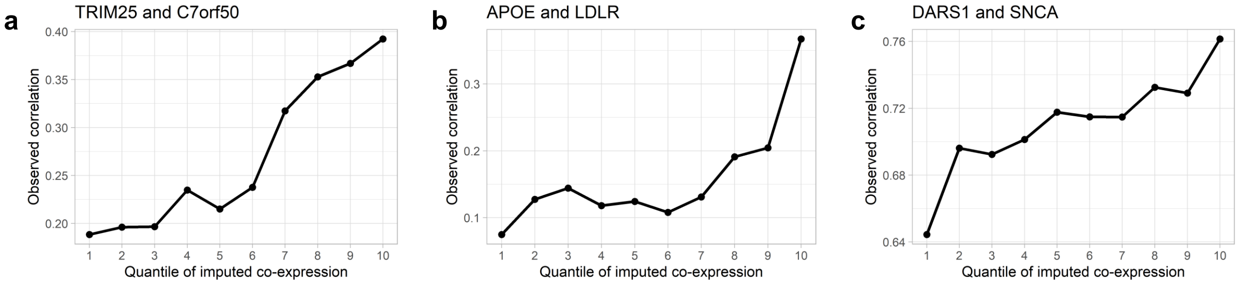


**Figure S1: Relationship between observed correlation and imputed co-expression.** **a**, **b**, **c**, Observed correlation of protein expression within each decile of imputed co-expression. The protein pairs shown are TRIM25 and C7orf50 (**a**), APOE and LDLR (**b**), and DARS1 and SNCA (**c**). These three protein pairs were arbitrarily selected from among the pairs identified as significant by the COWAS interaction test for low-density lipoprotein (LDL) cholesterol, Alzheimer's disease, and Parkinson's disease, respectively.

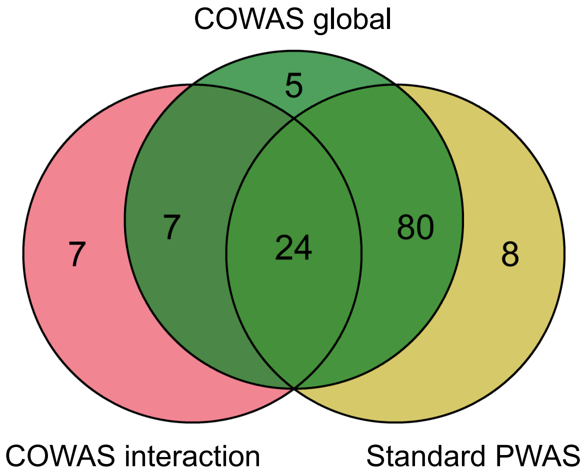


**Figure S2: Numbers of significant protein pairs for LDL cholesterol.**
A Venn diagram displaying the numbers of protein pairs identified as significant for low-density lipoprotein (LDL) cholesterol by the COWAS global test (green), the COWAS interaction test (pink), or a standard PWAS analysis (yellow). Here "standard PWAS" refers to pairs in which at least one of the proteins was identified by PWAS.

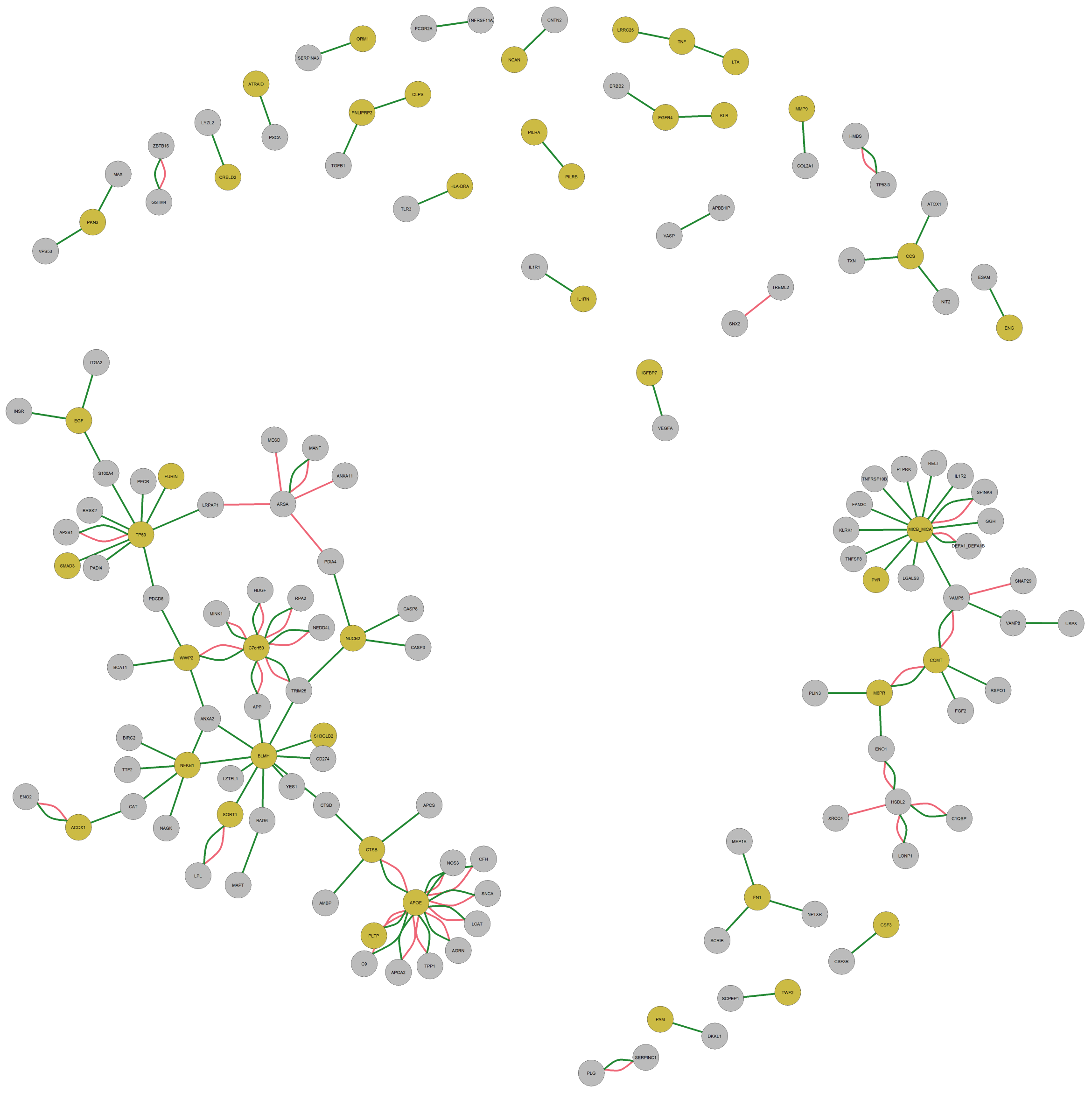


**Figure S3: Network of COWAS discoveries for LDL cholesterol.**
A network diagram showing all of the protein pairs identified as significant for low-density lipoprotein (LDL) cholesterol by either the COWAS global test (green edges) or the COWAS interaction test (pink edges). Node colors indicate whether each protein was identified as significant for LDL cholesterol by standard PWAS (yellow) or not (gray).

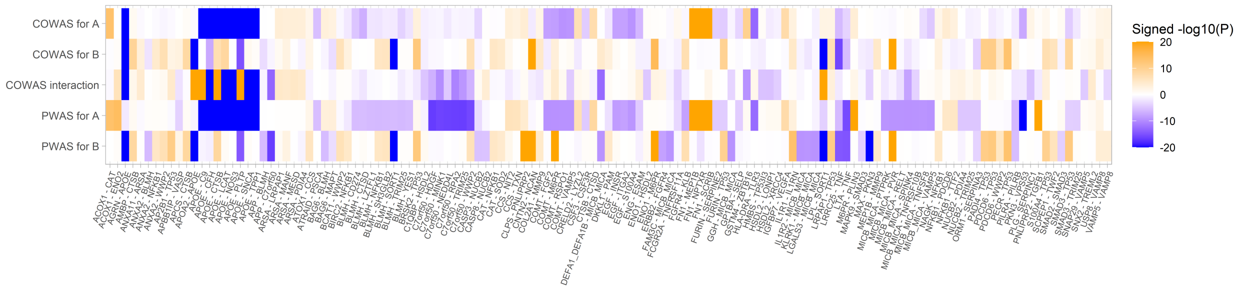


**Figure S4: $P$ values and effect directions for LDL cholesterol.**
A heat map displaying COWAS single-protein and interaction test results, as well as standard PWAS results, for low-density lipoprotein (LDL) cholesterol levels. Orange denotes positive effects and blue denotes negative effects, with color intensity corresponding to the $-\log_{10}(P)$ value of the test. All protein pairs identified as significant by the COWAS global test, the COWAS interaction test, or a standard PWAS analysis are included in this figure. From top to bottom, the rows correspond to single-protein COWAS tests for proteins A and B, the COWAS interaction test, and PWAS tests for proteins A and B. Here A and B refer to the first and second proteins listed in each pair, respectively. To facilitate visualization, the $-\log_{10}(P)$ values were capped at 20.

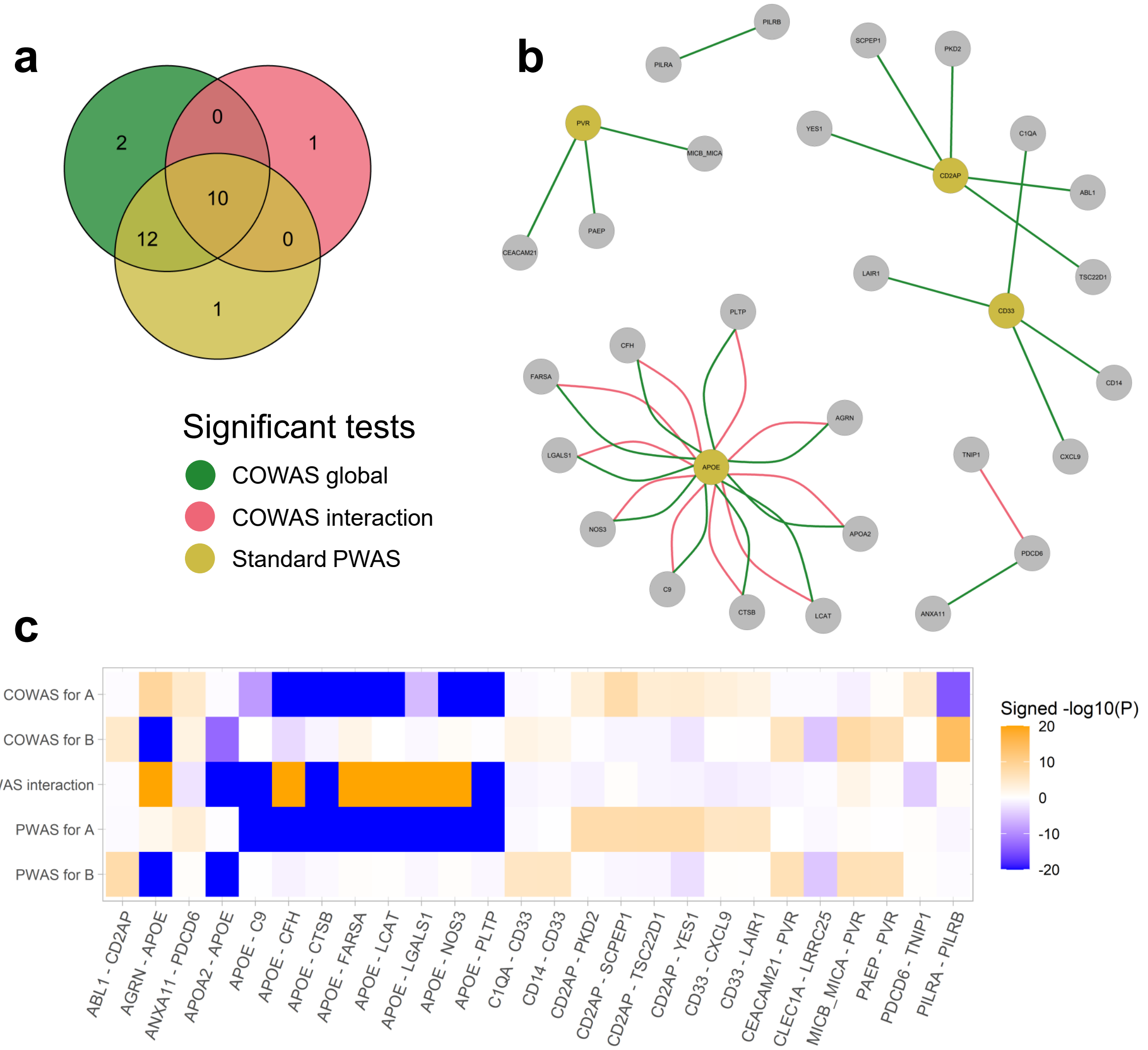


**Figure S5: COWAS and PWAS results for Alzheimer's disease on IGAP GWAS data.** **a**, A Venn diagram displaying the numbers of protein pairs identified as significant for Alzheimer's disease (AD) using IGAP GWAS data by the COWAS global test (green), the COWAS interaction test (pink), or a standard PWAS analysis (yellow). Here "standard PWAS" refers to pairs in which at least one of the proteins was identified by PWAS. **b**, A network diagram showing all of the protein pairs identified as significant for AD using IGAP GWAS data by either the COWAS global test (green edges) or the COWAS interaction test (pink edges). Node colors indicate whether each protein was identified as significant for AD by standard PWAS using IGAP GWAS data (yellow) or not (gray). **c**, A heat map displaying COWAS single-protein and interaction test results, as well as standard PWAS results, for all protein pairs identified as significant for AD using IGAP GWAS data by at least one of the three tests shown in **a**. Orange denotes positive effects and blue denotes negative effects, with color intensity corresponding to the $-\log_{10}(P)$ value of the test. A and B refer to the first and second proteins listed in each pair, respectively. To facilitate visualization, the $-\log_{10}(P)$ values were capped at 20.

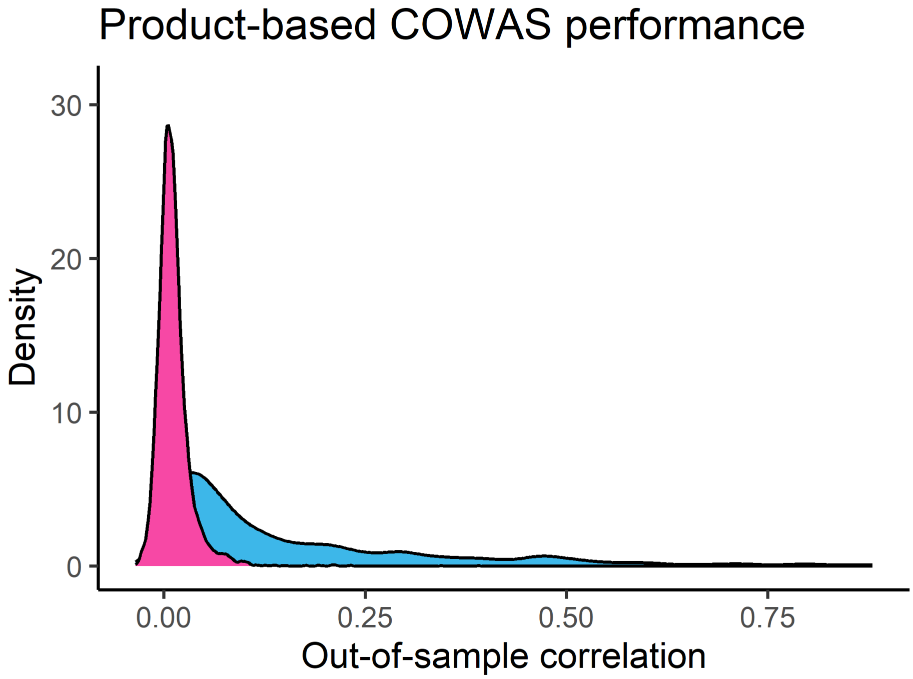


**Figure S6: Predictive performance of product-based COWAS imputation models.** A density plot displaying out-of-sample correlations for product-based COWAS models. Correlation between the observed product of expression levels and its prediction is shown in pink, while correlation between observed single-protein expression and its prediction is shown in blue. Imputation models were trained using elastic net linear regression with the top 100 *cis*-SNPs pre-selected according to their pQTL effect sizes. Only variants present in the EADB GWAS of Alzheimer's disease were used for model training.

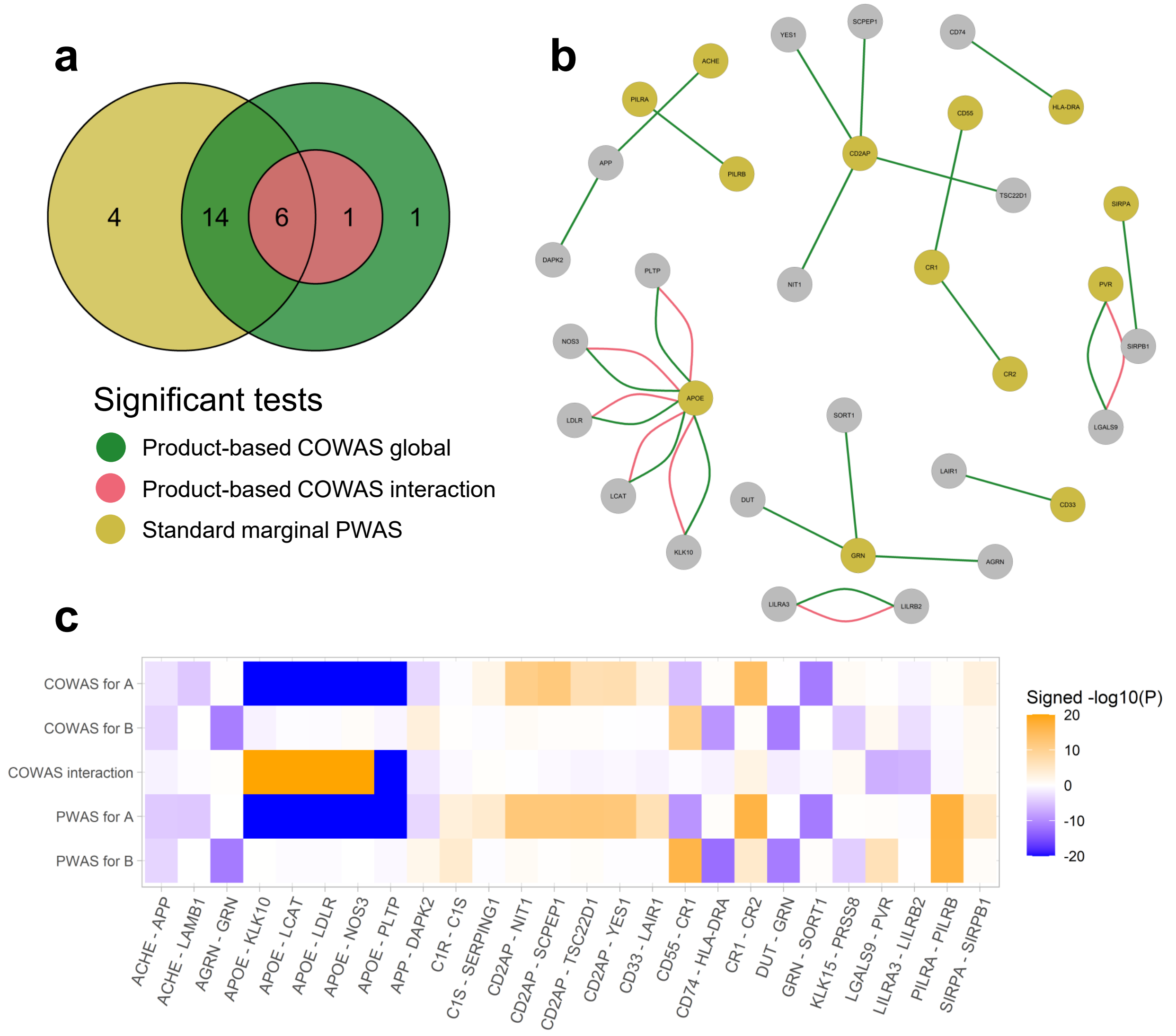


**Figure S7: Product-based COWAS and PWAS results for Alzheimer's disease.** **a**, A Venn diagram displaying the numbers of protein pairs identified as significant for Alzheimer's disease (AD) using EADB GWAS data by the product-based COWAS global test (green), the product-based COWAS interaction test (pink), or a standard PWAS analysis (yellow). Here "standard PWAS" refers to pairs in which at least one of the proteins was identified by PWAS. **b**, A network diagram showing all of the protein pairs identified as significant for AD using EADB GWAS data by either the product-based COWAS global test (green edges) or the product-based COWAS interaction test (pink edges). Node colors indicate whether each protein was identified as significant for AD by standard PWAS using EADB GWAS data (yellow) or not (gray). **c**, A heat map displaying product-based COWAS single-protein and interaction test results, as well as standard PWAS results, for all protein pairs identified as significant for AD using EADB GWAS data by at least one of the three tests shown in **a**. Orange denotes positive effects and blue denotes negative effects, with color intensity corresponding to the $-\log_{10}(P)$ value of the test. A and B refer to the first and second proteins listed in each pair, respectively. To facilitate visualization, the $-\log_{10}(P)$ values were capped at 20.

### 2 Supplementary tables

##### Table S1: Significant COWAS and PWAS test results for Alzheimer's disease using EADB GWAS summary statistics

| Proteins A - B | COWAS for A | | COWAS for B | | COWAS interaction | | COWAS global | PWAS for A | | PWAS for B | |
|---|---|---|---|---|---|---|---|---|---|---|---|
| | Effect (SE) | *P* | Effect (SE) | *P* | Effect (SE) | *P* | *P* | Effect (SE) | *P* | Effect (SE) | *P* |
| APOE - LDLR | -0.19 (0.003) | **0.00** | -0.02 (0.022) | 0.46 | 0.42 (0.021) | **3.6e-89** | **0.00** | -0.17 (0.003) | **0.00** | -0.02 (0.023) | 0.48 |
| APP - DAPK2 | -0.06 (0.019) | 8.0e-04 | 0.05 (0.014) | 4.1e-04 | -0.29 (0.102) | 4.8e-03 | **1.2e-05** | -0.06 (0.019) | 7.7e-04 | 0.01 (0.005) | 0.01 |
| ARHGEF10 - CD2AP | 8.7e-04 (0.005) | 0.87 | 0.05 (0.008) | **3.6e-09** | -0.04 (0.058) | 0.51 | **2.0e-11** | -2.4e-04 (0.005) | 0.96 | 0.05 (0.007) | **4.7e-13** |
| C1QA - CD33 | 2.2e-03 (0.004) | 0.62 | 0.02 (0.005) | 1.08e-03 | -0.40 (0.221) | 0.07 | **1.4e-06** | 2.0e-03 (0.004) | 0.65 | 0.01 (0.002) | **2.7e-07** |
| CD14 - CD33 | -0.01 (0.007) | 0.24 | 0.02 (0.005) | 2.3e-03 | -0.33 (0.224) | 0.14 | **1.4e-06** | -0.01 (0.007) | 0.28 | 0.01 (0.002) | **2.5e-07** |
| CD2AP - PKD2 | 0.04 (0.008) | **1.8e-06** | 0.01 (0.017) | 0.41 | -0.33 (0.170) | 0.05 | **2.3e-12** | 0.05 (0.007) | **3.7e-13** | 0.01 (0.017) | 0.39 |
| CD2AP - TSC22D1 | 0.04 (0.008) | **3.4e-07** | 0.04 (0.036) | 0.31 | -0.12 (0.072) | 0.09 | **3.5e-12** | 0.05 (0.007) | **4.7e-13** | 0.04 (0.036) | 0.29 |
| CD33 - CXCL9 | 0.02 (0.005) | **6.9e-05** | 0.01 (0.015) | 0.66 | -0.78 (0.311) | 0.01 | **2.4e-07** | 0.01 (0.002) | **2.7e-07** | 0.01 (0.015) | 0.36 |
| CD33 - LAIR1 | 0.02 (0.006) | 6.3e-04 | -1.6e-03 (0.003) | 0.61 | -0.26 (0.124) | 0.04 | **8.3e-07** | 0.01 (0.002) | **2.6e-07** | -1.1e-03 (0.003) | 0.73 |
| CD33 - LEP | 0.02 (0.006) | 7.2e-04 | 0.06 (0.060) | 0.29 | -0.81 (0.395) | 0.04 | **5.8e-07** | 0.01 (0.002) | **2.7e-07** | 0.07 (0.060) | 0.28 |
| CDH1 - SIRPA | 0.04 (0.036) | 0.26 | 0.01 (0.006) | 0.04 | -0.14 (0.144) | 0.35 | 2.0e-04 | 0.04 (0.036) | 0.27 | 0.01 (0.002) | **2.8e-05** |
| CNTN2 - CNTNAP2 | -0.01 (0.004) | 9.6e-04 | -1.8e-03 (0.004) | 0.67 | -0.31 (0.080) | **8.4e-05** | 1.0e-03 | -2.0e-04 (0.002) | 0.94 | 3.4e-03 (0.004) | 0.38 |
| ELOA - SIRPA | -0.01 (0.021) | 0.48 | 0.01 (0.005) | 0.02 | -0.42 (0.490) | 0.39 | 3.1e-04 | -0.01 (0.021) | 0.50 | 0.01 (0.002) | **2.9e-05** |
| GRN - PDCD6 | -0.06 (0.009) | **1.1e-11** | 1.6e-03 (0.003) | 0.61 | -0.08 (0.078) | 0.27 | **7.0e-11** | -0.06 (0.009) | **2.8e-12** | -9.1e-04 (0.002) | 0.65 |
| IGF1R - SIRPA | -0.02 (0.013) | 0.15 | 0.01 (0.007) | 0.13 | -0.12 (0.208) | 0.56 | 1.9e-04 | -0.02 (0.013) | 0.16 | 0.01 (0.002) | **2.9e-05** |
| LILRB2 - NOTCH1 | -0.02 (0.005) | 1.6e-04 | 0.02 (0.015) | 0.13 | 0.63 (0.157) | **5.7e-05** | 3.4e-04 | -5.1e-04 (0.002) | 0.80 | 0.02 (0.015) | 0.13 |
| MICB_MICA - PVR | -2.5e-03 (0.003) | 0.46 | 0.01 (0.003) | **3.5e-06** | 3.1e-03 (0.099) | 0.98 | **4.1e-06** | -2.4e-03 (0.002) | 0.15 | 0.01 (0.002) | **4.2e-07** |
| PILRA - PILRB | 0.04 (0.044) | 0.36 | -0.02 (0.038) | 0.60 | 0.02 (0.050) | 0.67 | **4.0e-16** | 0.02 (0.002) | **6.6e-18** | 0.02 (0.002) | **9.4e-18** |
| SIGLEC7 - TLR3 | 0.02 (0.005) | **2.8e-05** | 1.8e-03 (0.003) | 0.56 | -0.01 (0.078) | 0.91 | 3.5e-04 | 0.02 (0.005) | **2.7e-05** | 1.4e-03 (0.002) | 0.38 |
| SIGLEC9 - TLR3 | 0.01 (0.003) | **1.4e-05** | 1.2e-03 (0.003) | 0.64 | 0.02 (0.156) | 0.89 | 2.1e-04 | 0.01 (0.003) | **1.5e-05** | 1.4e-03 (0.002) | 0.38 |
| SIRPA - SIRPB1 | 0.01 (0.007) | 0.15 | 2.8e-03 (0.002) | 0.26 | -0.08 (0.176) | 0.65 | 2.7e-04 | 0.01 (0.002) | **3.0e-05** | 2.9e-03 (0.002) | 0.24 |

##### Table S2: Significant COWAS and PWAS test results for Alzheimer's disease using IGAP GWAS summary statistics

| Proteins A - B | COWAS for A | | COWAS for B | | COWAS interaction | | COWAS global | PWAS for A | | PWAS for B | |
|---|---|---|---|---|---|---|---|---|---|---|---|
| | Effect (SE) | *P* | Effect (SE) | *P* | Effect (SE) | *P* | *P* | Effect (SE) | *P* | Effect (SE) | *P* |
| ABL1 - CD2AP | -0.05 (0.061) | 0.45 | 0.09 (0.022) | **2.3e-05** | -0.12 (0.176) | 0.50 | **5.2e-07** | -0.05 (0.061) | 0.39 | 0.10 (0.018) | **2.7e-08** |
| AGRN - APOE | 0.08 (0.014) | **1.2e-09** | -0.24 (0.010) | **2.1e-132** | 4.45 (0.244) | **3.4e-74** | **0.00** | 0.03 (0.014) | 0.02 | -0.37 (0.006) | **0.00** |
| ANXA11 - PDCD6 | 0.24 (0.057) | **2.7e-05** | 0.06 (0.021) | 2.7e-03 | -0.63 (0.221) | 4.4e-03 | **5.8e-05** | 0.20 (0.055) | 2.7e-04 | 0.01 (0.006) | 0.35 |
| APOA2 - APOE | -0.04 (0.065) | 0.50 | -0.10 (0.013) | **9.1e-14** | -4.28 (0.184) | **3.7e-119** | **0.00** | -0.03 (0.067) | 0.63 | -0.37 (0.006) | **0.00** |
| APOE - C9 | -0.08 (0.014) | **2.1e-09** | 0.01 (0.032) | 0.79 | -5.91 (0.243) | **4.6e-131** | **0.00** | -0.37 (0.006) | **0.00** | -0.01 (0.033) | 0.79 |
| APOE - CFH | -0.26 (0.009) | **7.6e-191** | -0.05 (0.014) | 5.6e-04 | 2.12 (0.113) | **3.4e-79** | **0.00** | -0.37 (0.006) | **0.00** | -0.03 (0.015) | 0.08 |
| APOE - CTSB | -0.31 (0.007) | **0.00** | -0.01 (0.009) | 0.13 | -4.10 (0.227) | **1.2e-72** | **0.00** | -0.37 (0.006) | **0.00** | -0.01 (0.009) | 0.34 |
| APOE - FARSA | -0.21 (0.011) | **4.0e-90** | 0.17 (0.064) | 0.01 | 7.76 (0.419) | **1.0e-76** | **0.00** | -0.37 (0.006) | **0.00** | 0.05 (0.066) | 0.46 |
| APOE - LCAT | -0.29 (0.008) | **5.3e-255** | 6.9e-04 (0.037) | 0.99 | 1.94 (0.120) | **2.7e-58** | **0.00** | -0.37 (0.006) | **0.00** | 0.02 (0.038) | 0.64 |
| APOE - LGALS1 | -0.07 (0.014) | **1.3e-06** | -0.02 (0.018) | 0.24 | 12.42 (0.516) | **7.3e-128** | **0.00** | -0.37 (0.006) | **0.00** | -0.02 (0.019) | 0.23 |
| APOE - NOS3 | -0.23 (0.010) | **6.5e-122** | 0.10 (0.074) | 0.20 | 4.23 (0.215) | **3.3e-86** | **0.00** | -0.38 (0.006) | **0.00** | 0.06 (0.077) | 0.42 |
| APOE - PLTP | -0.19 (0.010) | **2.0e-76** | -0.01 (0.013) | 0.64 | -2.29 (0.106) | **1.2e-103** | **0.00** | -0.37 (0.006) | **0.00** | -0.02 (0.013) | 0.22 |
| C1QA - CD33 | -0.01 (0.012) | 0.31 | 0.04 (0.015) | 0.01 | -0.91 (0.602) | 0.13 | **2.1e-05** | -0.01 (0.012) | 0.29 | 0.02 (0.005) | **4.4e-06** |
| CD14 - CD33 | -0.01 (0.019) | 0.73 | 0.03 (0.014) | 0.02 | -0.54 (0.561) | 0.34 | **6.2e-05** | -0.01 (0.019) | 0.79 | 0.02 (0.005) | **4.3e-06** |
| CD2AP - PKD2 | 0.08 (0.022) | 5.1e-04 | -0.02 (0.047) | 0.73 | -0.73 (0.405) | 0.07 | **2.0e-07** | 0.10 (0.018) | **3.1e-08** | -0.02 (0.047) | 0.74 |
| CD2AP - SCPEP1 | 0.11 (0.019) | **2.6e-08** | -0.02 (0.015) | 0.13 | 0.07 (0.100) | 0.48 | **3.3e-07** | 0.10 (0.018) | **3.1e-08** | -0.02 (0.014) | 0.19 |
| CD2AP - TSC22D1 | 0.08 (0.022) | 1.2e-04 | -0.15 (0.100) | 0.15 | -0.29 (0.198) | 0.14 | **1.1e-07** | 0.10 (0.018) | **2.5e-08** | -0.14 (0.100) | 0.15 |
| CD2AP - YES1 | 0.08 (0.021) | **6.8e-05** | -0.13 (0.046) | 0.01 | -0.22 (0.131) | 0.09 | **1.7e-09** | 0.10 (0.018) | **2.7e-08** | -0.14 (0.045) | 1.5e-03 |
| CD33 - CXCL9 | 0.06 (0.017) | 6.4e-04 | 0.01 (0.043) | 0.83 | -2.40 (1.058) | 0.02 | **7.2e-06** | 0.02 (0.005) | **4.4e-06** | 0.03 (0.042) | 0.55 |
| CD33 - LAIR1 | 0.05 (0.018) | 3.0e-03 | 2.4e-03 (0.009) | 0.78 | -0.69 (0.371) | 0.06 | **1.7e-05** | 0.02 (0.005) | **4.3e-06** | 3.3e-03 (0.009) | 0.70 |
| CEACAM21 - PVR | -0.01 (0.006) | 0.39 | 0.03 (0.007) | **2.2e-06** | -1.13 (0.843) | 0.18 | **4.1e-06** | 3.9e-04 (0.005) | 0.94 | 0.03 (0.007) | **3.4e-07** |
| CLEC1A - LRRC25 | -0.01 (0.015) | 0.37 | -0.08 (0.017) | **1.2e-05** | -0.26 (0.319) | 0.42 | 1.1e-04 | -0.02 (0.014) | 0.26 | -0.07 (0.017) | **1.3e-05** |
| MICB_MICA - PVR | -0.02 (0.009) | 0.06 | 0.04 (0.007) | **9.0e-09** | 0.62 (0.236) | 0.01 | **2.4e-07** | 3.3e-03 (0.005) | 0.47 | 0.03 (0.007) | **3.0e-07** |
| PAEP - PVR | 0.02 (0.024) | 0.32 | 0.03 (0.007) | **4.0e-07** | 0.76 (0.684) | 0.27 | **5.6e-06** | -1.4e-03 (0.008) | 0.85 | 0.03 (0.007) | **3.7e-07** |
| PDCD6 - TNIP1 | 0.09 (0.021) | **3.6e-05** | 0.04 (0.073) | 0.62 | -0.98 (0.241) | **4.6e-05** | 5.6e-04 | 4.3e-03 (0.005) | 0.44 | 0.04 (0.073) | 0.59 |
| PILRA - PILRB | -0.64 (0.081) | **2.5e-15** | 0.54 (0.069) | **4.1e-15** | 0.17 (0.148) | 0.24 | **1.1e-13** | -0.01 (0.006) | 0.18 | -3.8e-03 (0.005) | 0.47 |

#### Table S3: Significant product-based COWAS and PWAS test results for Alzheimer's disease using EADB GWAS summary statistics

| Proteins A - B | COWAS for A | | COWAS for B | | COWAS interaction | | COWAS global | PWAS for A | | PWAS for B | |
|---|---|---|---|---|---|---|---|---|---|---|---|
| | Effect (SE) | *P* | Effect (SE) | *P* | Effect (SE) | *P* | *P* | Effect (SE) | *P* | Effect (SE) | *P* |
| ACHE - APP | -0.02 (0.005) | 3.3e-03 | -0.07 (0.019) | 2.3e-04 | -0.20 (0.114) | 0.07 | **1.5e-07** | -0.02 (0.005) | **2.5e-05** | -0.07 (0.019) | 2.5e-04 |
| ACHE - LAMB1 | -0.02 (0.005) | **1.6e-05** | 4.4e-04 (0.003) | 0.88 | -0.06 (0.117) | 0.61 | 2.9e-04 | -0.02 (0.005) | **1.6e-05** | 1.4e-04 (0.003) | 0.96 |
| AGRN - GRN | 3.7e-03 (0.007) | 0.60 | -0.06 (0.009) | **7.1e-12** | 0.11 (0.180) | 0.55 | **1.5e-10** | 5.5e-04 (0.005) | 0.91 | -0.07 (0.009) | **3.6e-12** |
| APOE - KLK10 | -0.19 (0.003) | **0.00** | -0.01 (0.003) | 0.08 | 0.93 (0.046) | **1.6e-88** | **0.00** | -0.17 (0.003) | **0.00** | 5.1e-04 (0.003) | 0.86 |
| APOE - LCAT | -0.17 (0.003) | **0.00** | -0.01 (0.014) | 0.54 | 0.73 (0.048) | **5.4e-53** | **0.00** | -0.17 (0.003) | **0.00** | -0.01 (0.014) | 0.48 |
| APOE - LDLR | -0.21 (0.004) | **0.00** | -0.02 (0.024) | 0.45 | 0.14 (0.008) | **5.7e-71** | **0.00** | -0.17 (0.003) | **0.00** | -0.02 (0.024) | 0.46 |
| APOE - NOS3 | -0.19 (0.003) | **0.00** | 0.03 (0.026) | 0.29 | 0.86 (0.046) | **7.7e-78** | **0.00** | -0.17 (0.003) | **0.00** | 2.0e-04 (0.026) | 0.99 |
| APOE - PLTP | -0.16 (0.003) | **0.00** | -0.01 (0.005) | 0.15 | -0.34 (0.035) | **7.0e-22** | **0.00** | -0.17 (0.003) | **0.00** | -0.01 (0.005) | 0.25 |
| APP - DAPK2 | -0.07 (0.020) | 4.4e-04 | 0.05 (0.014) | 6.9e-04 | -0.29 (0.109) | 0.01 | **1.2e-05** | -0.07 (0.020) | 4.3e-04 | 0.01 (0.005) | 0.02 |
| C1R - C1S | -0.01 (0.017) | 0.53 | 0.01 (0.023) | 0.69 | -0.06 (0.062) | 0.32 | 3.4e-04 | 0.02 (0.006) | 6.1e-04 | 0.02 (0.004) | **4.5e-05** |
| C1S - SERPING1 | 0.02 (0.009) | 0.01 | -4.1e-03 (0.006) | 0.52 | 0.05 (0.077) | 0.48 | 5.4e-04 | 0.02 (0.004) | **4.5e-05** | -4.0e-03 (0.006) | 0.52 |
| CD2AP - NIT1 | 0.05 (0.007) | **1.6e-11** | 0.01 (0.008) | 0.15 | -0.01 (0.037) | 0.79 | **1.3e-11** | 0.05 (0.007) | **7.2e-13** | 0.01 (0.008) | 0.13 |
| CD2AP - SCPEP1 | 0.05 (0.007) | **1.5e-12** | 4.5e-03 (0.006) | 0.43 | -0.04 (0.038) | 0.31 | **2.1e-11** | 0.05 (0.007) | **7.1e-13** | 2.3e-03 (0.005) | 0.65 |
| CD2AP - TSC22D1 | 0.04 (0.008) | **1.4e-07** | 0.02 (0.024) | 0.38 | -0.11 (0.070) | 0.11 | **4.3e-12** | 0.05 (0.007) | **4.6e-13** | 0.02 (0.024) | 0.31 |
| CD2AP - YES1 | 0.04 (0.008) | **7.2e-08** | -0.01 (0.017) | 0.75 | -0.08 (0.063) | 0.22 | **2.2e-11** | 0.05 (0.007) | **9.8e-13** | -0.01 (0.017) | 0.64 |
| CD33 - LAIR1 | 0.02 (0.006) | 1.7e-03 | -1.2e-03 (0.003) | 0.71 | -0.26 (0.151) | 0.08 | **1.7e-06** | 0.01 (0.002) | **2.7e-07** | -1.1e-03 (0.003) | 0.73 |
| CD55 - CR1 | -0.02 (0.005) | **2.5e-06** | 0.03 (0.004) | **7.2e-11** | -0.02 (0.021) | 0.43 | **1.4e-20** | -0.03 (0.005) | **5.7e-10** | 0.03 (0.004) | **5.2e-17** |
| CD74 - HLA-DRA | 0.02 (0.025) | 0.38 | -0.02 (0.003) | **7.7e-10** | -0.06 (0.032) | 0.09 | **2.3e-12** | 0.02 (0.025) | 0.38 | -0.02 (0.003) | **2.7e-13** |
| CR1 - CR2 | 0.03 (0.004) | **1.1e-14** | 0.01 (0.009) | 0.31 | 0.13 (0.045) | 3.7e-03 | **1.7e-17** | 0.03 (0.004) | **2.9e-17** | 0.03 (0.008) | **2.5e-05** |
| DUT - GRN | -0.01 (0.031) | 0.85 | -0.06 (0.009) | **4.8e-12** | -0.16 (0.070) | 0.03 | **2.2e-11** | 4.5e-04 (0.031) | 0.99 | -0.06 (0.009) | **4.6e-12** |
| GRN - SORT1 | -0.06 (0.009) | **3.5e-12** | 1.9e-03 (0.012) | 0.88 | -0.06 (0.054) | 0.29 | **7.4e-11** | -0.06 (0.009) | **3.6e-12** | 0.01 (0.010) | 0.40 |
| KLK15 - PRSS8 | 3.3e-03 (0.002) | 0.16 | -0.06 (0.016) | **3.9e-05** | 0.19 (0.112) | 0.09 | 2.5e-04 | 1.4e-03 (0.002) | 0.49 | -0.06 (0.015) | **6.3e-05** |
| LGALS9 - PVR | 1.7e-03 (0.004) | 0.70 | 0.01 (0.003) | 0.06 | -0.34 (0.065) | **1.3e-07** | **9.5e-12** | 3.1e-03 (0.004) | 0.47 | 0.01 (0.002) | **3.6e-07** |
| LILRA3 - LILRB2 | -3.7e-03 (0.002) | 0.13 | -0.01 (0.004) | 1.5e-03 | -0.03 (0.005) | **2.9e-07** | **6.3e-06** | -1.2e-03 (0.002) | 0.52 | -3.7e-04 (0.002) | 0.86 |
| PILRA - PILRB | 0.04 (0.044) | 0.32 | -0.03 (0.040) | 0.41 | -0.03 (0.026) | 0.27 | **2.4e-16** | 0.02 (0.002) | **6.7e-18** | 0.02 (0.002) | **9.4e-18** |
| SIRPA - SIRPB1 | 0.01 (0.002) | 1.2e-03 | 0.01 (0.003) | 0.05 | 0.05 (0.025) | 0.08 | **6.7e-05** | 0.01 (0.002) | **3.1e-05** | 2.9e-03 (0.002) | 0.24 |

#### Table S4: Significant COWAS and PWAS test results for Parkinson's disease

| Proteins A - B | COWAS for A | | COWAS for B | | COWAS interaction | | COWAS global | PWAS for A | | PWAS for B | |
|---|---|---|---|---|---|---|---|---|---|---|---|
| | Effect (SE) | *P* | Effect (SE) | *P* | Effect (SE) | *P* | *P* | Effect (SE) | *P* | Effect (SE) | *P* |
| AP1G2 - HLA-DRA | -0.02 (0.010) | 0.07 | -0.01 (0.003) | **4.3e-06** | -0.16 (0.107) | 0.14 | **3.4e-05** | -0.01 (0.009) | 0.14 | -0.01 (0.003) | **1.3e-05** |
| DARS1 - SNCA | 0.01 (0.025) | 0.80 | -0.04 (0.024) | 0.07 | 0.29 (0.039) | **1.2e-13** | **7.9e-14** | 3.8e-03 (0.025) | 0.88 | -0.07 (0.024) | 2.6e-03 |
| ENSA - SNCA | -0.03 (0.034) | 0.39 | -0.06 (0.028) | 0.02 | 0.27 (0.047) | **1.0e-08** | **2.1e-09** | -0.02 (0.034) | 0.54 | -0.09 (0.028) | 1.4e-03 |
| GRK5 - SNCA | -0.07 (0.026) | 4.3e-03 | -0.06 (0.022) | 0.01 | 0.31 (0.086) | 2.7e-04 | **4.8e-06** | 0.02 (0.008) | 0.04 | -0.07 (0.022) | 1.7e-03 |
| HCLS1 - SNCA | 0.03 (0.022) | 0.22 | -0.06 (0.029) | 0.02 | 0.14 (0.034) | **5.5e-05** | **1.2e-06** | 0.03 (0.022) | 0.21 | -0.10 (0.028) | 4.1e-04 |
| HIP1R - TRIM25 | 0.10 (0.020) | **6.2e-07** | 0.01 (0.018) | 0.72 | 0.17 (0.094) | 0.07 | **8.2e-06** | 0.09 (0.020) | **1.6e-06** | 0.01 (0.018) | 0.74 |
| SNCA - USP8 | -0.05 (0.024) | 0.06 | 0.06 (0.030) | 0.06 | 0.56 (0.109) | **3.6e-07** | **9.8e-09** | -0.08 (0.024) | 9.0e-04 | 0.05 (0.030) | 0.07 |

### 3 Supplementary notes

#### Note 1 Definition of product-based COWAS

As described in the main text, the interaction model in the main version of COWAS predicts the product of expression residuals, after removing the component of expression explained by linear genetic effects on its mean. We also considered modeling co-expression by predicting the product of observed expression levels. Here we define this alternative version of our method, which we refer to as product-based COWAS to distinguish it from the main version of our method, which we refer to as residual-based COWAS. Our implementation of COWAS in R (https://github.com/mykmal/cowas) allows users to choose whether to train models predicting an interaction between single-exposure model residuals (residual-based COWAS) or an interaction between observed expression levels (product-based COWAS).

Following the same notation defined in the main text, let $A_i$ and $B_i$ be real-valued random variables that represent the expression or abundance levels of two molecular exposures for some individual $i$. Further, let $\boldsymbol{Z}_{A,i}$ be a random vector of length $p_A$ whose entries contain dosage values in individual $i$ for the $p_A$ xQTLs that regulate the first exposure. Similarly, let $\boldsymbol{Z}_{B,i}$ be a random vector of length $p_B$ containing dosage values in individual $i$ for the $p_B$ xQTLs that regulate the second exposure. Finally, let $Y_i$ be a real-valued random variable representing the value of the outcome trait in individual $i$.

In stage 1 of product-based COWAS, we assume the following single-protein models:

$$A_i = \gamma_A + \boldsymbol{Z}_{A,i}^\intercal \boldsymbol{\beta}_A + \varepsilon_{A,i}, \tag{1}$$

$$B_i = \gamma_B + \boldsymbol{Z}_{B,i}^\intercal \boldsymbol{\beta}_B + \varepsilon_{B,i}. \tag{2}$$

Recall that in the residual-based version of COWAS we defined $C_i = \varepsilon_{A,i} \cdot \varepsilon_{B,i}$ and then assumed that $C_i$ itself also depends on genetic variation. Now, instead of modeling co-expression as the conditional expectation of that product of error terms, we will instead model it as the conditional expectation of the interaction between the centered expression values $A_i - E(A_i)$ and $B_i - E(B_i)$.

Let $T_i = \big(A_i - E(A_i)\big)\big(B_i - E(B_i)\big)$ denote the product of the centered expression levels of the two exposures. We assume that this product can be modeled as a linear combination of xQTL genotypes. Thus, we obtain the following model:

$$T_i = \gamma_T + \boldsymbol{Z}_i^\intercal \boldsymbol{\beta}_T + \varepsilon_{T,i}. \tag{3}$$

Here $\boldsymbol{Z}_i \in \mathbb{R}^p$ is a random vector whose entries contain dosage values in individual $i$ for all $p$ xQTLs that regulate the two exposures. The parameter $\gamma_T \in \mathbb{R}$ is the true intercept term, $\boldsymbol{\beta}_T \in \mathbb{R}^p$ is the true vector of genetic variant weights, and $\varepsilon_{T,i}$ is a real-valued random variable with mean zero.

In the second stage of product-based COWAS, we model the effect of the genetic component of $T_i$ on the outcome trait $Y_i$ while again accounting for direct effects from $E(A_i \mid \boldsymbol{Z}_{A,i})$ and $E(B_i \mid \boldsymbol{Z}_{B,i})$. Formally, our model for the outcome trait is

$$Y_i = \gamma_{YT} + E(A_i \mid \boldsymbol{Z}_{A,i})\theta_{AT} + E(B_i \mid \boldsymbol{Z}_{B,i})\theta_{BT} + E(T_i \mid \boldsymbol{Z}_i)\theta_T + \varepsilon_{YT,i}. \tag{4}$$

Here $\gamma_{YT} \in \mathbb{R}$ is the true intercept, $\theta_{AT}, \theta_{BT}, \theta_T \in \mathbb{R}$ are the true effect sizes, and the error term $\varepsilon_{YT,i}$ is a real-valued random variable with mean zero. Note that we used different subscripts here to distinguish these parameters from the parameters in the residual-based formulation of COWAS.

Two-sample model estimation can be performed similarly to how it was described for residual-based COWAS. Again following the notation defined in the main text, suppose we have two individual-level datasets sampled from the same population with sample sizes $n_1$ and $n_2$. Let

$\boldsymbol{A}, \boldsymbol{B} \in \mathbb{R}^{n_1}$ be observed vectors of the expression or abundance levels of the two exposures, as measured in the first dataset. Also let $\mathbf{Z}_{\text{A}} \in \mathbb{R}^{n_1 \times p_A}$ be the observed genotype matrix of $p_A$ xQTLs for the first exposure, and let $\mathbf{Z}_{\text{B}} \in \mathbb{R}^{n_1 \times p_B}$ be the observed genotype matrix of $p_B$ xQTLs for the second exposure, as genotyped in the same dataset of $n_1$ individuals. Then let $\mathbf{Z} \in \mathbb{R}^{n_1 \times p}$ be the observed joint genotype matrix of all $p$ xQTLs. Analogously we define $\mathbf{Z}^*_{\text{A}} \in \mathbb{R}^{n_2 \times p_A}$, $\mathbf{Z}^*_{\text{B}} \in \mathbb{R}^{n_2 \times p_B}$, and $\mathbf{Z}^* \in \mathbb{R}^{n_2 \times p}$ for the second dataset. Finally, let $\boldsymbol{Y}^* \in \mathbb{R}^{n_2}$ be an observed vector of outcome trait measurements for each individual in the second dataset. All of these vectors and each column of these matrices are assumed to be centered around zero and scaled to have a variance of one.

In the model training stage of product-based COWAS (stage 1), we first regress $\boldsymbol{A}$ and $\boldsymbol{B}$ on $\mathbf{Z}_{\text{A}}$ and $\mathbf{Z}_{\text{B}}$, respectively. This yields estimates of the parameter vectors $\boldsymbol{\beta}_A$ and $\boldsymbol{\beta}_B$. Then we compute $\boldsymbol{T} = \boldsymbol{A} \odot \boldsymbol{B}$ as the Hadamard product of $\boldsymbol{A}$ and $\boldsymbol{B}$. No additional centering of $\boldsymbol{A}$ and $\boldsymbol{B}$ is needed because we assumed that they were already standardized to have mean zero. Next, we regress $\boldsymbol{T}$ on $\mathbf{Z}$ to obtain an estimate of the parameter vector $\boldsymbol{\beta}_T$. In this paper, we used penalized linear regression to estimate the variant weights.

In the association testing stage of product-based COWAS (stage 2), the fitted weights $\widehat{\boldsymbol{\beta}}_A$, $\widehat{\boldsymbol{\beta}}_B$, and $\widehat{\boldsymbol{\beta}}_T$ are used to impute expression and co-expression for the $n_2$ samples in the second dataset. That is, we compute

$$\widehat{\boldsymbol{A}}^* = \mathbf{Z}^*_{\text{A}} \widehat{\boldsymbol{\beta}}_A, \tag{5}$$

$$\widehat{\boldsymbol{B}}^* = \mathbf{Z}^*_{\text{B}} \widehat{\boldsymbol{\beta}}_B, \tag{6}$$

$$\widehat{\boldsymbol{T}}^* = \mathbf{Z}^* \widehat{\boldsymbol{\beta}}_T. \tag{7}$$

Lastly, the product-based version of COWAS fits a multiple linear regression model with $\boldsymbol{Y}^*$ as the outcome and $\widehat{\boldsymbol{A}}^*, \widehat{\boldsymbol{B}}^*, \widehat{\boldsymbol{T}}^*$ as predictors to obtain estimates of the parameters $\theta_{AT}$, $\theta_{BT}$, and $\theta_T$. All of the same hypothesis tests described in the main text can be performed using product-based COWAS as well. Estimation and inference using GWAS summary statistics can also be done in exactly the same way as described in Note 2 for the residual-based version of COWAS, except that $\widehat{\boldsymbol{\beta}}_{co}$ is replaced with $\widehat{\boldsymbol{\beta}}_T$.

#### Note 2 Extension of COWAS for use with GWAS summary data

In this note we extend the association testing stage of COWAS (stage 2) so that it can be performed using summary-level GWAS data. The formulas we derive here only require fitted weights for expression and co-expression imputation models, $Z$ scores from a GWAS of the outcome trait, and a linkage disequilibrium (LD) reference panel. Note that all COWAS results reported in this paper were obtained using the summary-level version of our method. Our derivations follow the approach originally introduced in the multivariate imaging-wide association study (MV-IWAS) method [1, 2], which was based on earlier methods for multi-variant association testing using GWAS summary statistics [3–5].

Let $\widehat{\boldsymbol{\beta}}_A, \widehat{\boldsymbol{\beta}}_B, \widehat{\boldsymbol{\beta}}_{co} \in \mathbb{R}^p$ be the trained COWAS model weights for molecular phenotypes A, B, and their co-expression, respectively. Note that, unlike in the individual-level formulation of our method, the dimensions of all three weight vectors must match. This can be ensured by padding $\widehat{\boldsymbol{\beta}}_A$ and $\widehat{\boldsymbol{\beta}}_B$ with zeros in those positions where they are missing a variant compared to $\widehat{\boldsymbol{\beta}}_{co}$. We will denote the joint matrix of all model weights by $\widehat{\boldsymbol{\upbeta}} = (\widehat{\boldsymbol{\beta}}_A, \widehat{\boldsymbol{\beta}}_B, \widehat{\boldsymbol{\beta}}_{co}) \in \mathbb{R}^{p \times 3}$.

Furthermore, let $\widehat{z}_1, \ldots, \widehat{z}_p \in \mathbb{R}$ be $Z$ scores from a GWAS study of the outcome trait of interest for the same set of $p$ genetic variants. We assume that the GWAS was conducted in a population of the same genetic ancestry as the population used to train COWAS model weights. Importantly, reference and effect alleles must be consistent between the GWAS summary statistics and the

COWAS weights. Our implementation of COWAS in `R` automatically checks for allele consistency, flips GWAS $Z$ scores when necessary, and removes variants that cannot be harmonized. Next, COWAS converts the GWAS $Z$ scores to pseudocorrelation estimates. This is done by relying on the monotonic relationship between $Z$ scores and correlations [6], leading to the following formula for the pseudocorrelation between each variant $j$ and the outcome trait:

$$\widehat{c}_j = \frac{\widehat{z}_j}{\sqrt{n' - 1 + \widehat{z}_j^2}}, \tag{8}$$

where $n'$ is the sample size of the GWAS cohort for the outcome trait.

Finally, let $\mathbf{G} \in \mathbb{R}^{m \times p}$ be a genotype matrix for $m$ individuals and the same set of $p$ variants included in the COWAS models. We assume that these $m$ individuals are also of the same genetic ancestry as those used to train COWAS model weights. Moreover, we assume that each column of $\mathbf{G}$ has been centered around zero and scaled to a variance of one. An LD reference panel represents correlations among genetic variants, so we can compute it from $\mathbf{G}$ as follows:

$$\widehat{\mathbf{D}} = \frac{1}{m}\mathbf{G}^\intercal\mathbf{G}. \tag{9}$$

Now we will derive an estimator for $\boldsymbol{\theta} = (\theta_A, \theta_B, \theta_{co})^\intercal$ in terms of the trained COWAS model weights $\widehat{\boldsymbol{\beta}}$, the variant-outcome pseudocorrelation vector $\widehat{\boldsymbol{c}} = (\widehat{c}_1, \ldots, \widehat{c}_p)^\intercal$, and the LD reference panel $\widehat{\mathbf{D}}$. Suppose that $\widehat{\mathbf{X}}^* = (\widehat{\boldsymbol{A}}^*, \widehat{\boldsymbol{B}}^*, \widehat{\boldsymbol{C}}^*) \in \mathbb{R}^{n_2 \times 3}$ is an individual-level matrix of imputed expression and co-expression in the second-stage dataset of $n_2$ samples. Also suppose we have the outcome trait vector $\boldsymbol{Y}^* \in \mathbb{R}^{n_2}$ and individual-level genotype data $\mathbf{Z}^* \in \mathbb{R}^{n_2 \times p}$ for all $p$ variants and all $n_2$ individuals in the second-stage dataset. We assume that $\boldsymbol{Y}^*$ has been centered around zero and scaled to a variance of one, and that each column of $\mathbf{Z}^*$ has likewise been centered and scaled. We will first derive $\widehat{\boldsymbol{\theta}}$ and $\widehat{Var}(\widehat{\boldsymbol{\theta}})$ in terms of $\widehat{\mathbf{X}}^*$, $\boldsymbol{Y}^*$, and $\mathbf{Z}^*$, and then we will redefine our estimators in terms of summary-level quantities.

Regressing $\boldsymbol{Y}^*$ on $\widehat{\mathbf{X}}^*$ yields the following ordinary least squares estimator:

$$\widehat{\boldsymbol{\theta}} = \left((\widehat{\mathbf{X}}^*)^\intercal\widehat{\mathbf{X}}^*\right)^{-1}(\widehat{\mathbf{X}}^*)^\intercal\boldsymbol{Y}^* \tag{10}$$

$$= \left((\mathbf{Z}^*\widehat{\boldsymbol{\beta}})^\intercal\mathbf{Z}^*\widehat{\boldsymbol{\beta}}\right)^{-1}(\mathbf{Z}^*\widehat{\boldsymbol{\beta}})^\intercal\boldsymbol{Y}^* \tag{11}$$

$$= \left(\widehat{\boldsymbol{\beta}}^\intercal\frac{\mathbf{Z}^{*\intercal}\mathbf{Z}^*}{n_2}\widehat{\boldsymbol{\beta}}\right)^{-1}\widehat{\boldsymbol{\beta}}^\intercal\frac{\mathbf{Z}^{*\intercal}\boldsymbol{Y}^*}{n_2}. \tag{12}$$

Observe that $\frac{\mathbf{Z}^{*\intercal}\mathbf{Z}^*}{n_2}$ is a matrix of correlations among the genetic variants in $\mathbf{Z}^*$, so we can estimate it with $\widehat{\mathbf{D}}$. Moreover, $\frac{\mathbf{Z}^{*\intercal}\boldsymbol{Y}^*}{n_2}$ is a vector of correlations between each variant and $\boldsymbol{Y}^*$, so we can estimate it with $\widehat{\boldsymbol{c}}$. Therefore, the effects of genetically regulated expression and co-expression on the outcome trait are jointly estimated by

$$\widehat{\boldsymbol{\theta}} \approx \left(\widehat{\boldsymbol{\beta}}^\intercal\widehat{\mathbf{D}}\widehat{\boldsymbol{\beta}}\right)^{-1}\widehat{\boldsymbol{\beta}}^\intercal\widehat{\boldsymbol{c}}. \tag{13}$$

Similarly, the variance of $\widehat{\boldsymbol{\theta}}$ can be estimated in terms of $\widehat{\boldsymbol{\theta}}$, $\widehat{\boldsymbol{\beta}}$, $\widehat{\mathbf{D}}$, and $\widehat{\boldsymbol{c}}$. The residual sum of squares after regressing $\boldsymbol{Y}^*$ on the design matrix $\widehat{\mathbf{X}}^*$ is obtained as follows:

$$
\begin{aligned}
RSS &= \|\boldsymbol{Y}^* - \widehat{\mathbf{X}}^*\widehat{\boldsymbol{\theta}}\|^2 && (14)\\
&= \boldsymbol{Y}^{*\mathsf{T}}\boldsymbol{Y}^* - 2\boldsymbol{Y}^{*\mathsf{T}}\widehat{\mathbf{X}}^*\widehat{\boldsymbol{\theta}} + \widehat{\boldsymbol{\theta}}^{\mathsf{T}}(\widehat{\mathbf{X}}^*)^{\mathsf{T}}\widehat{\mathbf{X}}^*\widehat{\boldsymbol{\theta}} && (15)\\
&= (n_2-1)\frac{\boldsymbol{Y}^{*\mathsf{T}}\boldsymbol{Y}^*}{n_2-1} - 2\boldsymbol{Y}^{*\mathsf{T}}\mathbf{Z}^*\widehat{\boldsymbol{\upbeta}}\widehat{\boldsymbol{\theta}} + \widehat{\boldsymbol{\theta}}^{\mathsf{T}}\widehat{\boldsymbol{\upbeta}}^{\mathsf{T}}\mathbf{Z}^{*\mathsf{T}}\mathbf{Z}^*\widehat{\boldsymbol{\upbeta}}\widehat{\boldsymbol{\theta}} && (16)\\
&= (n_2-1)\frac{\boldsymbol{Y}^{*\mathsf{T}}\boldsymbol{Y}^*}{n_2-1} - 2n_2\Big(\frac{\mathbf{Z}^{*\mathsf{T}}\boldsymbol{Y}^*}{n_2}\Big)^{\mathsf{T}}\widehat{\boldsymbol{\upbeta}}\widehat{\boldsymbol{\theta}} + n_2\widehat{\boldsymbol{\theta}}^{\mathsf{T}}\widehat{\boldsymbol{\upbeta}}^{\mathsf{T}}\frac{\mathbf{Z}^{*\mathsf{T}}\mathbf{Z}^*}{n_2}\widehat{\boldsymbol{\upbeta}}\widehat{\boldsymbol{\theta}}. && (17)
\end{aligned}
$$

Observe that $\frac{\boldsymbol{Y}^{*\mathsf{T}}\boldsymbol{Y}^*}{n_2-1} = 1$ because we assumed that $\boldsymbol{Y}^*$ was scaled to have a variance of one, $\frac{\mathbf{Z}^{*\mathsf{T}}\boldsymbol{Y}^*}{n_2}$ can be estimated by $\widehat{\boldsymbol{c}}$, $\frac{\mathbf{Z}^{*\mathsf{T}}\mathbf{Z}^*}{n_2}$ can be estimated by $\widehat{\mathbf{D}}$, and the leftover $n_2$ can be replaced by $n'$. Therefore, we estimate the residual sum of squares by

$$
RSS \approx n'\Big(1 - 2\widehat{\boldsymbol{c}}^{\mathsf{T}}\widehat{\boldsymbol{\upbeta}}\widehat{\boldsymbol{\theta}} + \widehat{\boldsymbol{\theta}}^{\mathsf{T}}\widehat{\boldsymbol{\upbeta}}^{\mathsf{T}}\widehat{\mathbf{D}}\widehat{\boldsymbol{\upbeta}}\widehat{\boldsymbol{\theta}}\Big) - 1. \tag{18}
$$

Finally, we estimate the variance of $\widehat{\boldsymbol{\theta}}$ by

$$
\begin{aligned}
\widehat{Var}(\widehat{\boldsymbol{\theta}}) &= \Big((\widehat{\mathbf{X}}^*)^{\mathsf{T}}\widehat{\mathbf{X}}^*\Big)^{-1}\frac{RSS}{n_2-4} && (19)\\
&\approx \Big(\widehat{\boldsymbol{\upbeta}}^{\mathsf{T}}\widehat{\mathbf{D}}\widehat{\boldsymbol{\upbeta}}\Big)^{-1}\frac{RSS}{n'(n'-4)}. && (20)
\end{aligned}
$$

Wald tests for $\theta_A$, $\theta_B$, and $\theta_{co}$ can be performed in the same way as described for individual-level data in the main text. An $F$ test of overall significance can also be performed using the formula derived in the main text for individual-level data, except that the sample size of the individual-level outcome cohort ($n_2$) should be replaced with the sample size of the GWAS cohort ($n'$).

#### Note 3 Derivation of standard PWAS

Throughout this paper we compared our proposed method with the marginal, single-exposure PWAS approach commonly used today. In this note we define the standard PWAS model, explain how it is estimated using individual-level data, and then derive a summary-level version of its association testing stage. Without loss of generality, we will use our notation for protein A to explain the PWAS association test.

As before, let $A_i$ denote a real-valued random variable that represents the expression or abundance levels of protein A in individual $i$. Further, let $\boldsymbol{Z}_{A,i}$ be a random vector of length $p_A$ whose entries contain dosage values in individual $i$ for the $p_A$ xQTLs that regulate this exposure, and let $Y_i$ be a real-valued random variable representing the outcome trait of interest in individual $i$. The stage 1 model in PWAS is the following:

$$
A_i = \gamma_A + \boldsymbol{Z}_{A,i}^{\mathsf{T}}\boldsymbol{\beta}_A + \varepsilon_{A,i}. \tag{21}
$$

Here $\gamma_A \in \mathbb{R}$ is the true intercept, $\boldsymbol{\beta}_A \in \mathbb{R}^{p_A}$ is the vector of true genetic variant weights, and $\varepsilon_{A,i}$ is a real-valued random variable with mean zero.

In stage 2, PWAS assumes the following model:

$$
Y_i = \gamma_{YA,i} + E(A_i \mid \boldsymbol{Z}_{A,i})\theta_{AM,i} + \varepsilon_{YA,i}. \tag{22}
$$

Here $\gamma_{YA} \in \mathbb{R}$ is the true intercept, $\theta_{AM} \in \mathbb{R}$ is the true marginal effect of genetically regulated expression on the outcome trait, and $\varepsilon_{YA,i}$ is another real-valued random variable with mean zero.

Importantly, note that the PWAS effect size $\theta_{AM}$ is distinct from the COWAS effect size $\theta_A$. Whereas $\theta_{AM}$ is the marginal effect of $E(A_i \mid \boldsymbol{Z}_{A,i})$ on $Y_i$, the COWAS coefficient $\theta_A$ is the effect of $E(A_i \mid \boldsymbol{Z}_{A,i})$ on $Y_i$ after accounting for the effects of $E(B_i \mid \boldsymbol{Z}_{B,i})$ and $E(C_i \mid \boldsymbol{Z}_i)$ on $Y_i$.

To train PWAS model weights we need an individual-level dataset with genotype data and protein expression measurements. Let $\boldsymbol{A} \in \mathbb{R}^{n_1}$ be an observed vector of the expression or abundance levels of protein A, as measured in the stage 1 dataset of $n_1$ individuals. Also let $\mathbf{Z}_{\mathrm{A}} \in \mathbb{R}^{n_1 \times p_A}$ be the observed genotype matrix of $p_A$ pQTLs for this protein, as genotyped in the same dataset of $n_1$ individuals. Like before, we assume that $\boldsymbol{A}$ has been centered around zero and scaled to a variance of one, and that every column of $\mathbf{Z}_{\mathrm{A}}$ is likewise centered and scaled. The PWAS imputation model weight vector $\widehat{\boldsymbol{\beta}}_A$ is estimated by regressing $\boldsymbol{A}$ on $\mathbf{Z}_{\mathrm{A}}$. Any regression model can be used for this purpose, such as the penalized linear regression models we utilized in this paper.

To estimate the second stage model in standard PWAS, first suppose that we have individual-level data available for the outcome trait. Let $\boldsymbol{Y}^* \in \mathbb{R}^{n_2}$ be an observed vector of outcome trait measurements in the stage 2 dataset, and let $\mathbf{Z}^*_{\mathrm{A}} \in \mathbb{R}^{n_2 \times p_A}$ be an observed genotype matrix for those same $n_2$ individuals and all $p_A$ genetic variants included in the stage 1 model. Once again, we assume that $\boldsymbol{Y}^*$ has been centered around zero and scaled to a variance of one, and that each column of $\mathbf{Z}^*_{\mathrm{A}}$ has likewise been centered and scaled. We first impute genetically regulated expression into the outcome trait dataset, obtaining

$$\widehat{\boldsymbol{A}}^* = \mathbf{Z}^*_{\mathrm{A}} \widehat{\boldsymbol{\beta}}_A. \tag{23}$$

Next, we fit a simple linear regression model with $\boldsymbol{Y}^*$ as the outcome and $\widehat{\boldsymbol{A}}^*$ as the predictor. This yields an estimate of the parameter $\theta_{AM}$.

To determine if the genetically regulated component of expression has a significant effect on the outcome trait, standard PWAS tests the hypothesis $H_0 : \theta_{AM} = 0$ against its two-sided alternative using a Wald test. The test statistic is $(\widehat{\theta}_{AM})^2 / Var(\widehat{\theta}_{AM})$, which asymptotically follows a $\chi^2$ distribution with one degree of freedom under $H_0$.

In practice, we performed the association testing stage of PWAS using summary-level GWAS data for the outcome trait and an LD reference panel. The derivations for summary-level PWAS are analogous to those for summary-level COWAS, so we skip the intermediate steps here. Let $\widehat{\boldsymbol{c}}_A = (\widehat{c}_1, \ldots, \widehat{c}_{p_A})^{\mathsf{T}}$ be a vector of pseudocorrelations between the outcome trait and each of the $p_A$ variants included in the stage 1 PWAS model. Also let $\widehat{\mathbf{D}}_{\mathrm{A}} \in \mathbb{R}^{p_A \times p_A}$ be an LD reference panel for those same $p_A$ genetic variants. Then we can estimate $\theta_{AM}$ by

$$\widehat{\theta}_{AM} \approx \left( \widehat{\boldsymbol{\beta}}_A^{\mathsf{T}} \widehat{\mathbf{D}}_{\mathrm{A}} \widehat{\boldsymbol{\beta}}_A \right)^{-1} \widehat{\boldsymbol{\beta}}_A^{\mathsf{T}} \widehat{\boldsymbol{c}}_A. \tag{24}$$

The residual sum of squares for the stage 2 PWAS model can be estimated by

$$RSS_A \approx n' \Big( 1 - 2 \widehat{\boldsymbol{c}}_A^{\mathsf{T}} \widehat{\boldsymbol{\beta}}_A \widehat{\theta}_{AM} + \widehat{\theta}_{AM}^{\mathsf{T}} \widehat{\boldsymbol{\beta}}_A^{\mathsf{T}} \widehat{\mathbf{D}}_{\mathrm{A}} \widehat{\boldsymbol{\beta}}_A \widehat{\theta}_{AM} \Big) - 1. \tag{25}$$

Finally, we can estimate the variance of $\widehat{\theta}_{AM}$ by

$$\widehat{Var}(\widehat{\theta}_{AM}) \approx \left( \widehat{\boldsymbol{\beta}}_A^{\mathsf{T}} \widehat{\mathbf{D}}_{\mathrm{A}} \widehat{\boldsymbol{\beta}}_A \right)^{-1} \frac{RSS_A}{n'(n'-2)}, \tag{26}$$

where $n'$ is the sample size of the outcome trait GWAS.
